# Federated analysis of the contribution of recessive coding variants to 29,745 developmental disorder patients from diverse populations

**DOI:** 10.1101/2023.07.24.23293070

**Authors:** V. Kartik Chundru, Zhancheng Zhang, Klaudia Walter, Sarah Lindsay, Petr Danecek, Ruth Y. Eberhardt, Eugene J. Gardner, Daniel S. Malawsky, Emilie M. Wigdor, Rebecca Torene, Kyle Retterer, Caroline F. Wright, Kirsty McWalter, Eamonn Sheridan, Helen V. Firth, Matthew E. Hurles, Kaitlin E. Samocha, Vincent D. Ustach, Hilary C. Martin

## Abstract

Autosomal recessive (AR) coding variants are a well-known cause of rare disorders. We quantified the contribution of these variants to developmental disorders (DDs) in the largest and most ancestrally diverse sample to date, comprising 29,745 trios from the Deciphering Developmental Disorders (DDD) study and the genetic diagnostics company GeneDx, of whom 20.4% have genetically-inferred non-European ancestries. The estimated fraction of patients attributable to exome-wide AR coding variants ranged from ∼2% to ∼18% across genetically-inferred ancestry groups, and was significantly correlated with the average autozygosity (r=0.99, p=5x10^-6^). Established AR DD-associated (ARDD) genes explained 90% of the total AR coding burden, and this was not significantly different between probands with genetically-inferred European versus non-European ancestries. Approximately half the burden in these established genes was explained by variants not already reported as pathogenic in ClinVar. We estimated that ∼1% of undiagnosed patients in both cohorts were attributable to damaging biallelic genotypes involving missense variants in established ARDD genes, highlighting the challenge in interpreting these. By testing for gene-specific enrichment of damaging biallelic genotypes, we identified two novel ARDD genes passing Bonferroni correction, *KBTBD2* (p=1x10^-7^) and *CRELD1* (p=9x10^-8^). Several other novel or recently-reported candidate genes were identified at a more lenient 5% false-discovery rate, including *ZDHHC16* and *HECTD4*. This study expands our understanding of the genetic architecture of DDs across diverse genetically-inferred ancestry groups and suggests that improving strategies for interpreting missense variants in known ARDD genes may allow us to diagnose more patients than discovering the remaining genes.

## Introduction

High-throughput exome and genome sequencing^1^ have revolutionised the diagnosis of developmental disorders (DD)^2^, typically allowing 30-40% of patients to obtain a genetic diagnosis^3, 4^. Multiple new DD-associated genes have been discovered by statistical analysis of sequence data from large, phenotypically heterogeneous cohorts^5–10^. For example, a recent study brought together >30,000 trios primarily from the Deciphering Developmental Disorders (DDD) study and the US-based diagnostic testing company GeneDx, and identified twenty-eight novel genes in which *de novo* mutations are likely to cause DDs^5^. This contrasts with the more traditional approach of phenotype-driven gene discovery based on small numbers of patients or families who appear to have the same rare, clinically recognizable disorder (e.g.^11, 12^), or the “Matchmaker Exchange” approach in which researchers identify additional patients from other cohorts who have potentially damaging variants in the same candidate disease gene as an index patient^13^.

The genetic architecture of DDs has been shown to vary between genetically-defined ancestry groups as a result of varying levels of consanguinity^6, 14^. In a study of 6,040 exome-sequenced patients from the DDD study, we previously estimated through exome-wide burden analysis that ∼4% of probands with European ancestries and ∼31% of those with Pakistani ancestries could be explained by autosomal recessive (AR) coding variants, versus ∼50% and ∼30% respectively explained by *de novo* coding mutations^6^. Forty-eight percent of the exome-wide burden of recessive causes was in known AR DD-associated (ARDD) genes, indicating that larger sample sizes would be required to find the additional genes. Here, we combine a larger set of DDD trios with data from GeneDx to study the recessive contribution to DDs in a set of 29,745 trio probands across twenty-two genetically-inferred ancestry (GIA) groups, of whom 20.4% have majority non-European ancestries. We first quantify the recessive contribution to DDs across GIA groups and the extent to which this is explained by known genes and known pathogenic variants. We then conduct gene-based burden testing to identify new recessive genes underlying DDs.

## Results

We analysed deidentified exome-sequence data from the DDD study (N=13,450 probands) and from GeneDx. Since the vast majority of DDD patients have at least one Human Phenotype Ontology (HPO) term under ‘abnormality of the nervous system’, we selected the 36,057 GeneDx patients with at least one such term for inclusion in this analysis. There were differences in the reported phenotype distributions between the cohorts (**Supplementary Figures 1 and 2; Supplementary Table 1**), but these are likely to be largely attributable to the way HPO terms were recorded: DDD clinicians recorded HPO terms that they thought were particularly distinctive and likely to be relevant to a monogenic disorder, whereas for GeneDx, the HPO terms were abstracted from each patient’s medical history (see Methods). Consequently, many of the terms that differed in prevalence between the cohorts were nonspecific and/or indicated common conditions (e.g. ‘failure to thrive’, ‘asthma’) (**Supplementary Note**). Both cohorts have considerable heterogeneity of phenotypic presentations and genetic etiologies, and a similar burden of *de novo* mutations^5^. Given all this, we decided that the two cohorts were sufficiently similar to combine them for the work in this paper.

**Figure 1.**
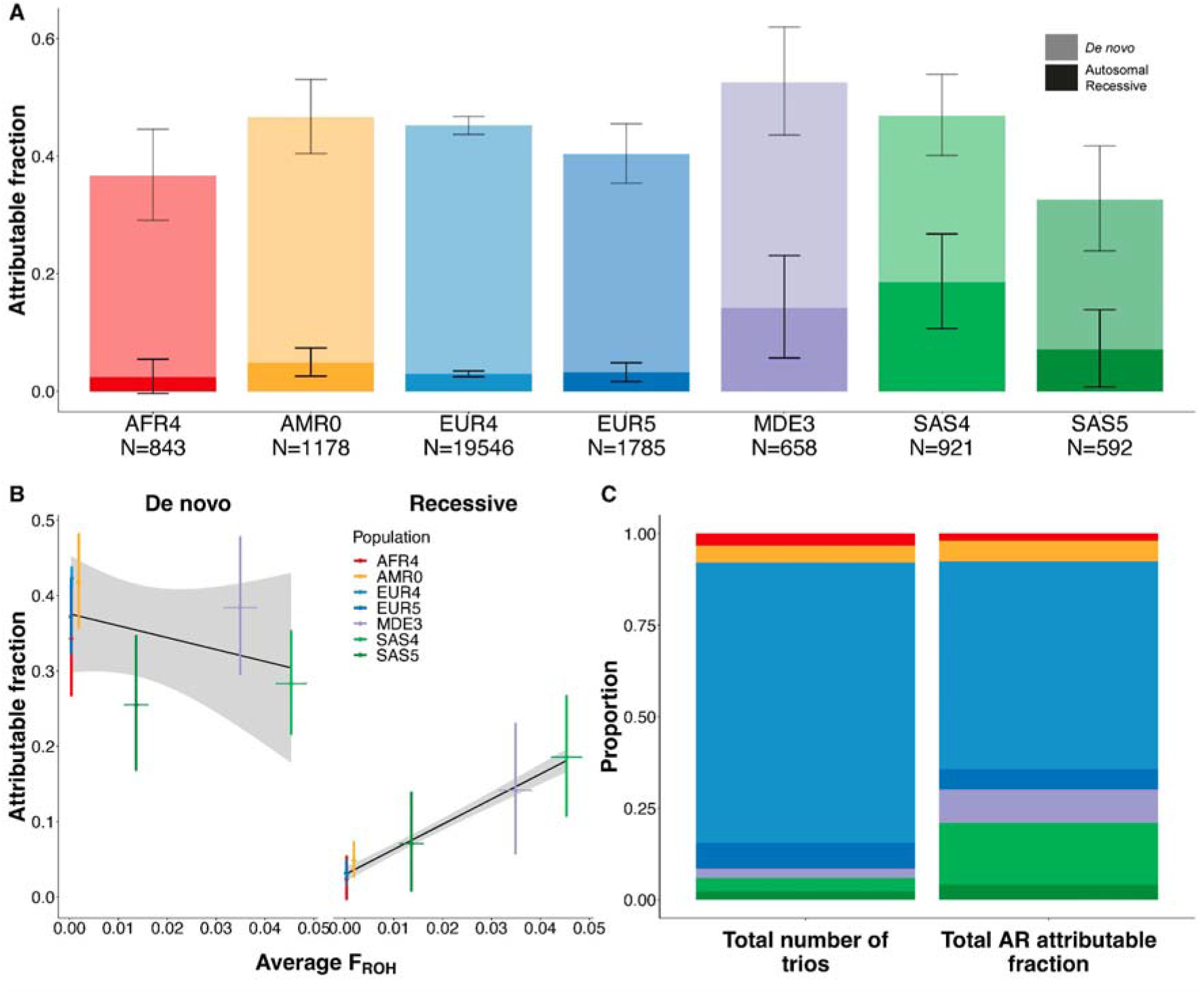
Estimates of the fraction of patients attributable to AR coding variants or *de novo* coding mutations in DDD and GeneDx across seven large GIA sub-groups. A) Estimated attributable fraction per GIA sub-group. The *de novo* attributable fractions (lighter shading) are stacked on the AR attributable fractions (darker shading), with the total height of the bars being the sum of the attributable fractions. Lines show 95% confidence intervals (CIs). B) Estimated attributable fraction due to *de novo* coding mutations (left) or AR coding variants (right) versus average autozygosity (F_ROH_) for these seven GIA sub-groups. Coloured lines show 95% CIs. The black line is the line of best fit, and grey shading shows its 95% confidence interval. C) Comparison of the proportion of the total sample size (left) versus the proportion of the total AR attributable fraction (right) accounted for by each GIA sub-groups.

**Figure 2:**
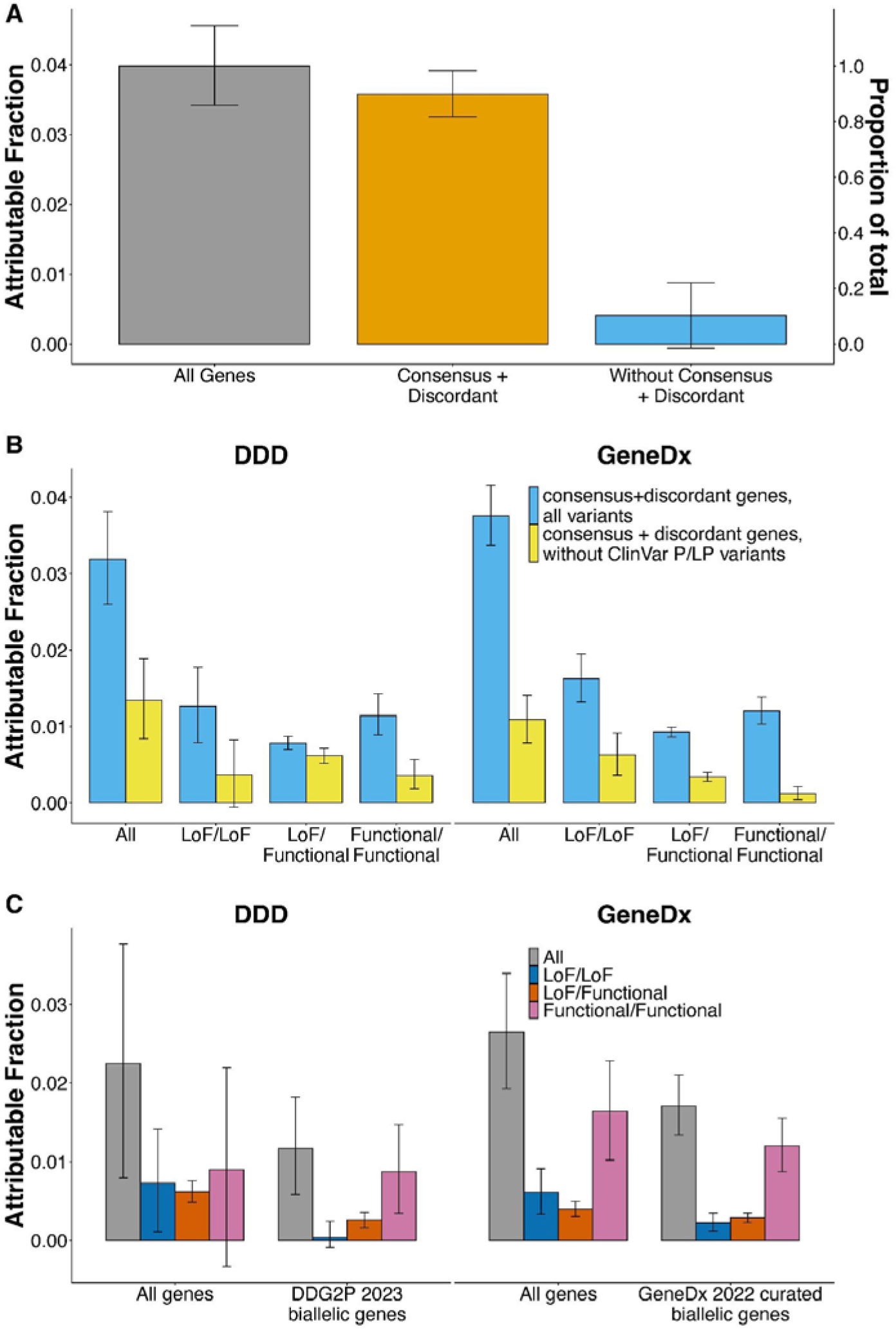
Estimates of the fraction of patients attributable to AR coding variants, in different subsets of genes and patients. These plots are focused on the individuals without cross-continental admixture from seven large GIA sub-groups, as in Figure 1 and Table 1. A) Estimates in all individuals from DDD and GeneDx combined, for all genes *versus* genes in the indicated lists. B) Estimates in all individuals for consensus+discordant genes split by cohort, comparing the estimates obtained with all variants *versus* after removing variants annotated as pathogenic/likely pathogenic in ClinVar. C) Estimates in undiagnosed individuals, for all genes *versus* the genes that are used for clinical filtering of diagnostic AR variants in the respective cohorts, split by cohort and functional consequence of the variants. Error bars show 95% confidence intervals.

**Table 1:** Sample sizes and average autozygosity values for the twenty-two GIA sub-groups included in the analyses, after removing probands with cross-continental admixture. The counts for all GIA sub-groups are shown in Supplementary Table 2. Note that DDD samples from these GIA sub-groups were excluded if there were fewer than a hundred unrelated, unaffected parents in DDD. *For some GIA sub-groups there were no reference samples in the same cluster (Supplementary Figure 4), so we give only the GIA groups label for these. ^+^ indicates the seven GIA sub-groups included in the overall attributable fraction calculations given throughout the text, in Figure 1 and in Supplementary Figures 11, 15 and 17. F_ROH_: fraction of the genome in runs of homozygosity (ROHs).

| GIA sub-group | Closest corresponding reference population | Number of unrelated probands | | | Number of unrelated, unaffected parents | | Average $F_{ROH}$ for probands | |
| --- | --- | --- | --- | --- | --- | --- | --- | --- |
|  |  | DDD | GeneDx | Combined | DDD | GeneDx | DDD | GeneDx |
| AFR3 | West African | - | 96 | 96 | - | 232 | - | 0.0013 |
| AFR4<sup>+</sup> | African-American | - | 843 | 843 | - | 2109 | - | 0.0003 |
| AMR0<sup>+</sup> | Mexican | - | 1178 | 1178 | - | 2674 | - | 0.0017 |
| AMR3 | Puerto Rican | - | 380 | 380 | - | 894 | - | 0.0027 |
| AMR4 | Latin American* | - | 204 | 204 | - | 511 | - | 0.0029 |
| AMR9 | Colombian | - | 260 | 260 | - | 745 | - | 0.0028 |
| EAS1 | East Asian* | - | 65 | 65 | - | 205 | - | 0.0003 |
| EAS2 | Chinese | - | 295 | 295 | - | 732 | - | 0.0008 |
| EAS3 | East Asian* | - | 128 | 128 | - | 348 | - | 0.0007 |
| EAS5 | Vietnamese/Cambodian | - | 89 | 89 | - | 245 | - | 0.0010 |
| EUR1 | European* | - | 657 | 657 | - | 1694 | - | 0.0021 |
| EUR3 | European* | - | 125 | 125 | - | 512 | - | 0.0005 |
| EUR4<sup>+</sup> | Western European | 7057 | 12489 | 19546 | 12997 | 26016 | 0.0005 | 0.0004 |
| EUR5<sup>+</sup> | Eastern European | 208 | 1577 | 1785 | 459 | 3909 | 0.0008 | 0.0002 |
| EUR6 | European* | - | 38 | 38 | - | 200 | - | 0.0001 |
| EUR7 | Italian | 118 | 1412 | 1530 | 294 | 3620 | 0.0010 | 0.0005 |
| MDE2 | Middle Eastern* | - | 79 | 79 | - | 149 | - | 0.0468 |
| MDE3<sup>+</sup> | Middle Eastern* | 87 | 571 | 658 | 157 | 1114 | 0.0351 | 0.0335 |
| MDE4 | Middle Eastern* | - | 87 | 87 | - | 167 | - | 0.0347 |
| SAS3 | Bangladeshi | 80 | 109 | 189 | 125 | 242 | 0.0185 | 0.0082 |
| SAS4<sup>+</sup> | Pakistani | 467 | 454 | 921 | 577 | 832 | 0.0539 | 0.0298 |
| SAS5<sup>+</sup> | Indian | 100 | 492 | 592 | 187 | 1019 | 0.0185 | 0.0108 |
| Total (all GIA sub-groups) |  | 8117 | 21628 | 29745 | 14796 | 48169 | 0.0044 | 0.0027 |
| Total (seven GIA sub-groups in Figure 1) |  | 7919 | 17604 | 25523 | 14377 | 37673 | 0.0043 | 0.0026 |

We began by classifying individuals into genetically-inferred ancestry (GIA) groups. The rationale for this was two-fold: firstly, we were interested in exploring differences in genetic architecture between these groups, and secondly, the analysis below relies on accurate estimates of allele frequencies which differ between groups. We recognize that these GIA groups do not capture the full genetic diversity of human populations. We determined the GIA groups for each cohort in a federated manner i.e. analysing summary statistics that were produced without physically combining the individual-level genetic data. The classifications were based on genetic similarity to individuals in the 1000 Genomes and Human Genome Diversity Panel (HGDP) reference datasets, inferred from principal component analysis (**Supplementary Figures 3** and **4**). We defined six continental-level GIA groups (AFR: African; AMR: Latin American; EAS: East Asian; EUR: European; MDE: Middle Eastern; SAS: South Asian) and, within these, forty-seven fine-scale GIA sub-groups (**Supplementary Table 2**).

We carried out federated quality control (QC) on the exome data across the two cohorts (**Supplementary Figures 5, 6** and **7)**, with sample QC done in a GIA-aware manner (**Supplementary Figure 8; Supplementary Table 3**) (see Methods). For the analyses described below, unless otherwise stated, we restricted to 29,745 unrelated trio probands from twenty-two GIA sub-groups (chosen as described in the Methods section “Sample filtering for the burden analysis”) for whom both parents were inferred to come from the same GIA group as the child, and from whom at least one parent was inferred to come from the same GIA sub-group as the child (**Table 1**; **Supplementary Table 2**).

### Exome-wide burden analysis

Following our previous work^6^, we calculated the expected probabilities of rare (minor allele frequency; MAF <0.01) biallelic genotypes (homozygous non-reference, or compound heterozygous genotypes) in each cohort and GIA sub-group separately, taking into account GIA sub-group-specific allele frequencies and autozygosity levels (**Supplementary Figure 9**). The four genotype consequence classes we considered included synonymous/synonymous (i.e. biallelic synonymous) as a negative control, plus three predicted damaging classes: LoF/LoF (loss-of-function), LoF/functional and functional/functional, where the “functional” class included protein-altering variants other than high-confidence LoFs that passed various deleteriousness filters (see Methods section on “Filtering of missense and other functional variants”).

To quantify the exome-wide recessive burden, we compared the expected number of biallelic genotypes in a given consequence class to the observed number. **Supplementary Figure 10** indicates that these generally agree well for biallelic synonymous genotypes in the largest GIA sub-groups, demonstrating that our QC is robust. For GIA sub-groups with smaller sample sizes, the observed number of biallelic synonymous genotypes was often significantly lower than expected (**Supplementary Table 4**), which we suspect is because our estimate of the expected number was being inflated due to overestimation of allele frequencies in the small sample sizes. Thus, for **Figure 1** and for the total estimates reported throughout the text and shown in **Figure 2** and **Supplementary Figures 11, 15 and 17**, we focused on seven large GIA sub-groups (see Methods section “Sample filtering for the burden analysis”): AFR4, AMR0, EUR4, EUR5, MDE3, SAS4 and SAS5 (total N=25,523 unrelated probands; **Table 1**). We used the observed and expected numbers of biallelic genotypes within these different GIA sub-groups to calculate the fraction of patients attributable to autosomal recessive coding causes, which we refer to below as the “attributable fraction”. Briefly, this was calculated as the difference between the observed and expected number of damaging biallelic genotypes divided by the number of probands (**Supplementary Table 4;** see Methods section “Testing for enrichment of biallelic genotypes over expectation”). We calculated the attributable fraction both exome-wide using all probands in different GIA sub-groups separately or in aggregate (**Figure 1**, **Supplementary Figures 12 and 17b**), as well as in different subsets of probands and gene sets (**Figure 2**; **Supplementary Figures 11 and 15**).

The estimates of attributable fraction due to AR coding variants ranged from ∼2-7% in AMR0, EUR4, EUR5 and SAS5 to 14.1% [95% CI: 5.6-23.0%] in MDE3 and 18.7% [10.8-26.9%] in SAS4. For all populations, this was lower than the attributable fraction due to *de novo* coding mutations (**Figure 1a**). The AR attributable fraction was significantly correlated with the average level of autozygosity in the GIA groups (r=0.99, p=5x10^-6^ for the 7 GIA sub-groups in **Figure 1b**), while the attributable fraction due to *de novo*s was not (r=-0.46, p=0.30; **Figure 1b**). Thus, despite making up only a small proportion of the total number of probands (6% combined), SAS4 and MDE3 make up 26.1% of the total AR attributable fraction across these seven GIA sub-groups (**Figure 1c**). **Supplementary Figure 11** shows that the total AR attributable fraction is slightly higher in GeneDx compared to DDD (4.4% versus 3.1%; p=1x10^-6^), which may reflect differences in the recruitment strategies for the two cohorts; as previously noted, DDD may be depleted of easy-to-diagnose recessive cases^6^. The recessive attributable fraction was also higher in diagnosed than undiagnosed patients (6.4% versus 2.6%; p=2.3x10^-49^), but similar in females and males (4.2% versus 3.8%; p=0.055) (**Supplementary Figure 11**).

We next examined how much of the exome-wide AR attributable fraction was explained by known disease-associated AR genes. We considered a set of 1,818 known ARDD genes that are used for diagnosis in one or both cohorts, including 1,069 “consensus” genes in both the DDG2P list and GeneDx’s in-house list, and 749 “discordant” genes in only one of the lists (**Supplementary Table 5**). Consensus genes explained 72.6% [65.6-79.8%] of the total exome-wide attributable fraction, and consensus+discordant genes explained 89.8% [81.7-98.3%] (**Figure 2a**). Once the consensus+discordant genes were removed, there was no significant residual burden of damaging biallelic genotypes across the remaining genes (attributable fraction=0.4% [-0.1-0.9%]; p=0.08) (**Figure 2a**). Consensus+discordant genes explained 91.0% [81.3%-101.1%] of the total exome-wide attributable fraction in probands with European ancestries (EUR4+EUR5), which was not significantly different from the fraction in those with non-European ancestries (AFR4+AMR0+MDE3+SAS4+SAS5) (87.9% [73.6%-103.0%]; p=0.9).

We removed variants annotated as pathogenic/likely pathogenic (P/LP) in ClinVar and estimated the remaining AR attributable fraction in the consensus+discordant ARDD genes. In DDD, we estimated that 42.2% [26.3-59.2%] of the AR attributable fraction in these genes was explained by variants not annotated as P/LP in ClinVar, but the fraction was a bit lower in GeneDx (29.1% [21.0-37.6%]) (**Figure 2b**), likely reflecting the fact that GeneDx systematically submits pathogenic variants to ClinVar whereas DDD does not. This implies that a substantial fraction of the recessive burden in the GIA groups represented by DDD is due to variants in known AR genes that have not been annotated as P/LP in ClinVar.

We then estimated the AR attributable fraction in as-yet-undiagnosed patients (N_undiagnosed_=4,425 and 12,604 for DDD and GeneDx respectively for the seven GIA-groups in **Figure 1**) within the set of ARDD genes that were used for diagnosis by the relevant cohort. From this, we estimated that 1.2% [0.6-1.8%] of the as-yet-undiagnosed DDD patients are attributable to damaging biallelic coding variants in AR DDG2P genes (**Figure 2c**). Almost all of this (96.8%) is due to biallelic LoF/functional or functional/functional genotypes. Amongst these 4,425 DDD undiagnosed individuals, there are 367 damaging biallelic genotypes in AR DDG2P genes that have been reported back to clinicians via DECIPHER as potentially clinically relevant and that have either not yet been clinically evaluated (106 genotypes), or have been classified as being of uncertain significance (261 genotypes). Thus, based on the above attributable fraction estimate, this implies that 14.1% ([1.168% of 4425]/367) of these biallelic genotypes are actually pathogenic. Similarly, in GeneDx, we estimate that 1.7% [1.3-2.1%] of as-yet-undiagnosed patients are attributable to damaging biallelic coding variants in known AR disease genes on GeneDx’s curated in-house list, of which 87.1% are biallelic LoF/functional or functional/functional. Many of these are likely being reported as VUSs as they do not currently meet the criteria for being classed as P/LP. This highlights the challenge of interpreting rare missense and other functional variants in ARDD genes.

Amongst the undiagnosed GeneDx patients, there was a significant excess of rare LoF/LoF genotypes in ARDD genes on the diagnostic list, and the attributable fraction due to such genotypes was estimated at 0.2% [0.1-0.3%] (**Figure 2c**). In contrast, there was no significant burden of LoF/LoF genotypes in AR DDG2P genes in undiagnosed DDD patients (attributable fraction 0.04% [-0.09-0.24%]; z-test for a difference in proportions compared to the GeneDx attributable fraction p=0.02). This may partly reflect the fact that GeneDx does not usually carry out reanalysis as new genes are added to their diagnostic list, unless it is requested by the clinician or as part of a research project, and there can be a lag in issuing updated reports for patients when a variant’s classification is upgraded from “variant of unknown significance” to “likely pathogenic”. In contrast, the iterative reanalysis carried out in DDD ensures that these relatively easy-to-interpret diagnostic variants in newly defined ARDD genes are not missed^15, 16^.

### Gene discovery

We next tested for an enrichment of damaging biallelic genotypes in each gene to try to identify novel ARDD genes. For the main gene discovery analysis, we included 29,745 unrelated trios without cross-continental admixture from the twenty-two GIA sub-groups shown in **Table 1**. We reasoned that although the expected number of biallelic genotypes was being overestimated in some of the smaller GIA sub-groups, this would only reduce our power rather than lead to false positives, and that the increased sample size by including these GIA sub-groups might compensate for this. For each gene, we used a Poisson test to compare the total observed number of biallelic genotypes in a given consequence class across GIA sub-groups with the total expected number (**Supplementary Figure 13**). We considered four combinations of damaging biallelic genotypes (LoF/LoF, LoF/LoF+LoF/functional, functional/functional, and all combined [LoF/LoF+LoF/functional+functional/functional]), and then took the lowest p-value per gene. We used a Bonferroni threshold of p < 7.2 x 10^-7^ (corrected for four tests per each of 17,320 genes).

We found twenty-five genes passing Bonferroni correction, and an additional forty-six genes passing a false-discovery rate (FDR) of 5% (**Supplementary Table 6**). Twenty-three of the Bonferroni significant genes (92%) and sixty-two of the FDR<5% genes (87.3%) are known ARDD genes on the GeneDx and/or DDG2P lists. **Table 2** shows the nine genes passing FDR<5% which are not on either list: *CRELD1*, *KBTBD2*, *ZDHHC16, ATG4C, HECTD4*, *ATAD2B, ATXN1, LRRC34,* and *C11ORF94*. Of these, *CRELD1* and *KBTBD2* pass Bonferroni correction. In **Supplementary Table 7**, we present the observed deleterious biallelic variants in these nine genes, together with the patients’ associated HPO terms. See the **Supplementary Note** for information on some of the suggestive genes.

**Table 2:**
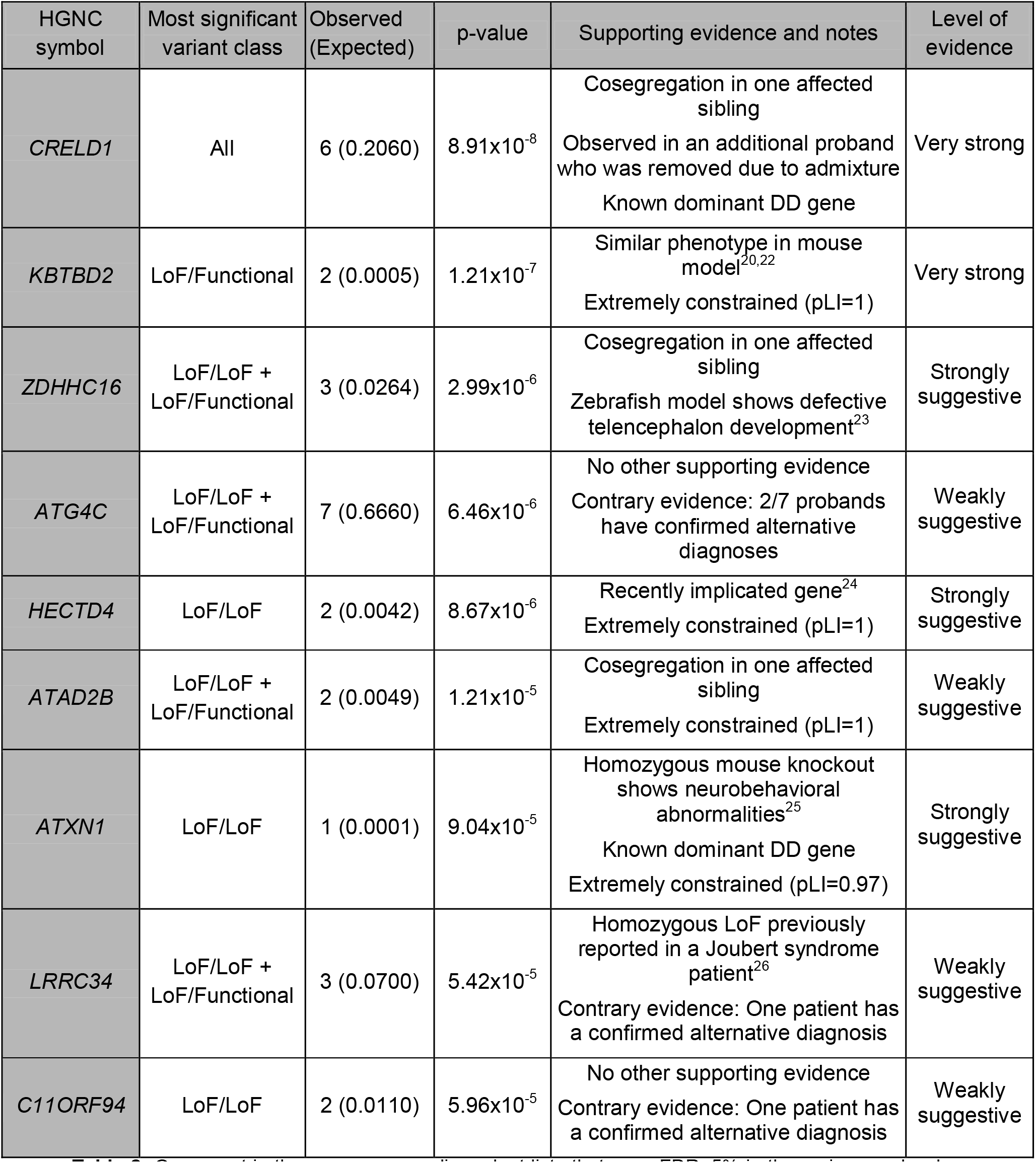
Genes not in the consensus or discordant lists that pass FDR<5% in the main gene burden analysis, which was based on 29,745 probands without inferred cross-continental admixture. P-values < 7.2x10^-7^ pass Bonferroni correction. We only show the result for the most significant combination of consequence classes per gene. **Supplementary Table 6** shows results for all combinations of consequence classes.

| HGNC symbol | Most significant variant class | Observed (Expected) | p-value | Supporting evidence and notes | Level of evidence |
| --- | --- | --- | --- | --- | --- |
| *CRELD1* | All | 6 (0.2060) | $8.91 \times 10^{-8}$ | Cosegregation in one affected sibling<br>Observed in an additional proband who was removed due to admixture<br>Known dominant DD gene | Very strong |
| *KBTBD2* | LoF/Functional | 2 (0.0005) | $1.21 \times 10^{-7}$ | Similar phenotype in mouse model<sup>20,22</sup><br>Extremely constrained (pLI=1) | Very strong |
| *ZDHHC16* | LoF/LoF + LoF/Functional | 3 (0.0264) | $2.99 \times 10^{-6}$ | Cosegregation in one affected sibling<br>Zebrafish model shows defective telencephalon development<sup>23</sup> | Strongly suggestive |
| *ATG4C* | LoF/LoF + LoF/Functional | 7 (0.6660) | $6.46 \times 10^{-6}$ | No other supporting evidence<br>Contrary evidence: 2/7 probands have confirmed alternative diagnoses | Weakly suggestive |
| *HECTD4* | LoF/LoF | 2 (0.0042) | $8.67 \times 10^{-6}$ | Recently implicated gene<sup>24</sup><br>Extremely constrained (pLI=1) | Strongly suggestive |
| *ATAD2B* | LoF/LoF + LoF/Functional | 2 (0.0049) | $1.21 \times 10^{-5}$ | Cosegregation in one affected sibling<br>Extremely constrained (pLI=1) | Weakly suggestive |
| *ATXN1* | LoF/LoF | 1 (0.0001) | $9.04 \times 10^{-5}$ | Homozygous mouse knockout shows neurobehavioral abnormalities<sup>25</sup><br>Known dominant DD gene<br>Extremely constrained (pLI=0.97) | Strongly suggestive |
| *LRRC34* | LoF/LoF + LoF/Functional | 3 (0.0700) | $5.42 \times 10^{-5}$ | Homozygous LoF previously reported in a Joubert syndrome patient<sup>26</sup><br>Contrary evidence: One patient has a confirmed alternative diagnosis | Weakly suggestive |
| *C11ORF94* | LoF/LoF | 2 (0.0110) | $5.96 \times 10^{-5}$ | No other supporting evidence<br>Contrary evidence: One patient has a confirmed alternative diagnosis | Weakly suggestive |

*CRELD1* (p=8.9x10^-8^) is an established monoallelic (dominant) DD gene on both the DDG2P and GeneDx lists, in which heterozygous missense mutations cause atrioventricular septal defect associated with heterotaxy syndrome^17, 18^. We observed eight probands with biallelic LoF/functional or functional/functional genotypes in this gene, all of whom had global developmental delay and seizures, plus variable other features. A concurrent study involving patients from GeneDx and other cohorts has also identified *CRELD1* as a novel ARDD gene with a differing phenotypic presentation to the dominant disorder it causes^19^. Three of the five GeneDx patients we identified were also in that study.

We observed two patients with damaging LoF/functional compound heterozygous genotypes in *KBTBD2* (p=1.2x10^-7^). The mouse knockout of this gene exhibits lipodystrophy, hepatic steatosis, insulin resistance, severe diabetes, and growth retardation^20^. Consistent with this, both patients displayed some degree of growth retardation. The older patient had hyperglycaemia and diabetes; the younger one was below the age at which diabetes might be expected to develop (**Supplementary Table 7**). Thus, the phenotypes in these patients appear to be consistent with the mouse knockout, but with some additional phenotypic features (microcephaly, cardiomyopathy, developmental delay).

We calculated semantic similarity scores^21^ of HPO terms between pairs of patients with damaging biallelic genotypes in the genes that passed FDR<5% correction, and compared the distribution of these scores to scores from randomly-chosen pairs of patients **(Supplementary Figure 14**). We found that the patients with damaging biallelic genotypes in the consensus+discordant genes were significantly more phenotypically similar to each other than were the randomly-chosen patients (one-sided Wilcoxon rank sum p=2.2x10^-126^), and also more similar than the patients with damaging biallelic genotypes in the nine novel FDR<5% genes (p=0.00376). However, the latter were also significantly more phenotypically similar to each other than were the randomly-chosen patients (p=0.024), which strengthens the evidence that they result in distinct phenotypes.

We repeated the gene-based tests using only undiagnosed probands (N=20,217 trios), after applying a stricter filter for admixed probands (requiring both parents to come from the same GIA sub-group as the child; N=23,574 trios), and after removing the filter of probands with inferred cross-continental admixture (N=32,058 trios). No additional novel genes were identified, and key conclusions were unchanged (**Supplementary Table 6**).

## Discussion

We have examined the contribution of AR coding variants to DDs in the largest sample to date, containing about six times more trios and greater ancestral diversity than our previous work in an earlier release of the DDD study^6^. The current study demonstrates the power of federated analysis of large multi-ancestry cohorts from which genome-wide individual-level data cannot be combined in a single location due to data governance considerations. Transferring deidentified summary data (principal components and their loadings) between cohorts allowed us to identify individuals with similar GIA across cohorts (**Supplementary Figure 4**), boosting power particularly for smaller and historically understudied groups. Using these multiple GIA groups, we showed that the average level of autozygosity is a strong predictor of the fraction of patients in a given sample who are attributable to AR causes (**Figure 1b**, **Supplementary Figure 12**).

We found that the great majority of the AR burden is explained by known ARDD genes, and that this is true both for European-ancestry and non-European-ancestry individuals (91% versus 88%). In contrast, our 2018 paper^6^ reported that amongst DDD probands with European and Pakistani GIA (roughly corresponding to the EUR4 and SAS4 GIA sub-groups in this work), the AR DDG2P genes known at the time explained 48% of the recessive burden. **Supplementary Figure 15** shows that the most recent DDG2P ARDD gene list explains a higher fraction of the burden in this analysis than the list used in our previous paper^6^, and that the GeneDx ARDD gene list explains the highest fraction. This suggests that DDG2P is a more conservatively curated list than the GeneDx list. It also reflects the success of worldwide AR gene-discovery efforts over the last five years, many of which have been conducted through MatchMaker Exchange-style approaches^13^. The comparably modest yield of new ARDD genes in this work suggests that this kind of statistical analysis of large, relatively unselected cohorts may not be the most efficient way to find new AR genes. A more efficient approach may entail identifying new candidate genes in unsolved cases in geographically-isolated populations or in consanguineous families with multiple affected individuals, then finding additional cases via Matchmaker Exchange or other global data sharing initiatives.

Our paper contains several clinically important messages. Firstly, the fact that most of the AR burden is in known ARDD genes suggests that, if a patient undergoes sequencing and is not found to have any candidate putatively damaging biallelic genotypes in these genes (of which only a subset meet ACMG P/LP criteria), their residual risk of having an AR condition, at least due to coding variants, is low. However, it does depend on their degree of consanguinity; from the attributable fraction estimates in our dataset, we estimate this residual risk at 0.43% [0.01-0.87%] if the patient has F_ROH_<0.0156 (the expectation for offspring of second cousins), and 14.56% [2.50-27.31-%] otherwise (F_ROH_>0.0156). Secondly, our estimates suggest that a substantial fraction of potential diagnoses in known AR genes are being missed, mostly those involving missense variants, which remain challenging to interpret (**Figure 2c**). For example, amongst the 7,732 DDD individuals included in our main burden analysis, there are 230 confirmed AR diagnoses in DDG2P genes, and the attributable fraction estimate in **Figure 2c** suggests that there are an additional ∼52 diagnoses to be made in these genes amongst the 4,425 undiagnosed patients (1.168% of 4,425); thus, we are missing ∼18.4% of diagnoses (52/282) in AR DDG2P genes. Thirdly, our results also imply that, if we could find all the possible diagnoses in established ARDD genes by better distinguishing pathogenic from benign functional variants, we would likely diagnose about twice as many patients as we would by discovering new ARDD genes, at least in the GIA groups under study here; our attributable fraction estimates suggest that, amongst undiagnosed DDD and GeneDx patients, there are ∼237 diagnoses yet to be made from LoF/functional and functional/functional genotypes in the ARDD genes that are already used for reporting by the relevant cohort, versus ∼103 diagnoses to be made from as-yet-undiscovered ARDD genes. Finally, the fact that ∼42% of the burden in established ARDD genes in DDD is not explained by ClinVar P/LP variants suggests that recessive carrier screening, which currently tends to focus on such variants^27^, has the potential to be extended in future as knowledge about which variants are clinically significant increases.

Despite the fact that the vast majority of the AR burden was explained by known genes in the GeneDx and/or DDG2P lists, we did identify several new or only recently-described genes with compelling or suggestive evidence for causation (**Table 2**). Overall, we consider *CRELD1* and *KBDBT2* to be confirmed *bona fide* ARDD genes, *ZDHHC16*, *HECTD4* and *ATXN1* to be highly suggestive (see **Supplementary Note**), and *ATG4C, ATAD2B, LRRC34* and *C11ORF94* to be weakly suggestive. Of the thirty-two damaging biallelic genotypes contributing to the discovery of these nine genes, twenty-seven were LoF/functional or functional/functional. Our stringent missense filtering (**Supplementary Figure 16** and **17**) boosted our power to implicate these genes: had we done more lenient missense filtering, the genes highlighted in **Table 2** would all have had less significant p-values, and none would have passed Bonferroni correction (**Supplementary Figure 18**).

This work has several limitations. Firstly, the families studied are not a random sample of the DD patient population, and may be depleted of easy-to-solve families with recessive conditions. Thus, we may have underestimated the contribution of AR variants to DDs as a whole, and hundreds of known ARDD genes did not reach formal Bonferonni significance in our study; we emphasise that the genes that passed Bonferroni correction in this work are not the only *bona fide* ARDD genes. Secondly, our estimates of attributable fraction assume that every excess damaging biallelic genotype over expectation fully ‘explains’ one proband (i.e. fully penetrant monogenic causes), which may over-simplify the genetic architecture. Thirdly, although our sample contained considerable ancestral diversity, it is clearly not representative of the global population, and the sample sizes for many GIA sub-groups were too small to get precise estimates of attributable fraction. This also undoubtedly reduced our power for gene discovery since parental allele frequencies are overestimated in these small samples. Finally, we focused on protein-coding single nucleotide variants and small indels, so our estimates do not include the recessive contribution of noncoding variants or copy number variants.

In conclusion, discovery of the remaining ARDD genes will require larger samples and/or more focused sampling of consanguineous families with multiple affected individuals. However, these as-yet-undiscovered genes are unlikely to account for a high fraction of patients, at least in the GIA groups represented in this study. To maximise diagnostic yield, future work should develop better strategies to distinguish pathogenic from benign functional recessive variants, such as approaches involving multiplex assays of variant effects^28, 29^.

### Data availability

Sequence and variant-level data and phenotype data from the DDD study data are available on the European Genome-phenome Archive (EGA; https://www.ebi.ac.uk/ega/) with study ID EGAS00001000775. GeneDx data cannot be made available through the EGA owing to the nature of consent for clinical testing. GeneDx-referred patients are consented for aggregate, deidentified research and subject to US HIPAA privacy protection. As such, we are not able to share patient-level BAM or VCF data, which are potentially identifiable without a HIPAA Business Associate Agreement. Access to the deidentified aggregate data used in this analysis is available upon request to GeneDx. GeneDx has contributed deidentified data to this study to improve clinical interpretation of genomic data, in accordance with patient consent and in conformance with the ACMG position statement on genomic data sharing. Clinically interpreted variants and associated phenotypes from the DDD study are available through DECIPHER (https://www.deciphergenomics.org/). Clinically interpreted variants from GeneDx are deposited in ClinVar (https://www.ncbi.nlm.nih.gov/clinvar) under accession number 26957 (https://www.ncbi.nlm.nih.gov/clinvar/submitters/26957/).

### Code availability

The code to perform the burden analysis and reproduce plots from this paper is available on GitHub (https://github.com/chundruv/DDD_GeneDx_Recessives), as is the code to run the Phenopy method (https://github.com/GeneDx/phenopy).

## Acknowledgements

VKC, ZZ, KW, SL, RT, VDU and HCM analysed the data. PD, RYE, EJG, DSM, EMW, and KS contributed to code, methods, and quality control analyses. RT, KR, CFW, MEH, KES, VDU and HCM supervised the study. HVF and ES provided clinical input. HCM conceived and directed the study. VKC and HCM wrote the first draft of the manuscript, with input from ZZ, KW and VDU. All authors commented on the final manuscript.

The DDD study presents independent research commissioned by the Health Innovation Challenge Fund (grant number HICF-1009-003). This study makes use of DECIPHER, which is funded by the Wellcome Trust. The full acknowledgements can be found at www.ddduk.org/access.html. We additionally thank Rachel Hobson, Erwan Delage and the Human Genetics Informatics team at Sanger for their input on the DDD study.

This research was funded in whole, or in part, by the Wellcome Trust Grant 220540/Z/20/A, ’Wellcome Sanger Institute Quinquennial Review 2021-2026’. For the purpose of Open Access, the author has applied a CC BY public copyright licence to any Author Accepted Manuscript version arising from this submission.

DSM is supported by a Gates Cambridge Scholarship (OPP1144).

## Methods

### Cohorts, sequencing, alignment and variant calling

#### Deciphering Developmental Disorders (DDD)

Between April 2011 and April 2015, the DDD study recruited a total of 13,450 patients (88% in a trio) with a severe developmental disorder who remained undiagnosed after undergoing the typical clinical genetics investigations^30^. The phenotypic inclusion criteria included neurodevelopmental disorders, congenital abnormalities, growth abnormalities, dysmorphic features, and unusual behavioural phenotypes. Recruitment took place across twenty-four regional genetics services within the United Kingdom and the Republic of Ireland health services. The families gave their informed consent to participate, and the study was approved by the UK Research Ethics Committee (10/H0305/83, granted by the Cambridge South Research Ethics Committee, and GEN/284/12, granted by the Republic of Ireland Research Ethics Committee).

Details on sample collection, exome sequencing, alignment, variant calling and variant annotation have been described previously^6, 7, 9^. Briefly, exome capture was carried out with either the Agilent SureSelect Human All Exon V3 or V5 baits. We used the BWA aln algorithm (BWA version 0.5.10) and the BWA mem algorithm (BWA version 0.7.12)^31, 32^ to align reads to the GRCh37 1000 Genomes Project phase 2 reference (hs37d5). Picard Markduplicates (versions 1.98 and 1.114)^33^ and Genome Analysis Toolkit IndelRealigner (GATK version 3.1.1 and version 3.5.0)^34–36^ were used for sample-level BAM improvement. To call single-nucleotide variants (SNVs) and indels, we used the GATK HaplotypeCaller, CombineGVCFs and GenotypeGVCFs (GATK version 3.5.0). Variant-calling was restricted to the merged bait regions from the Agilent V3 and V5 exome capture kits used in the sequencing, plus a padding region of 100 base pairs on either side.

#### GeneDx

Patients were referred to GeneDx for clinical whole-exome sequencing for diagnosis of suspected Mendelian disorders as previously described^5, 37^. Patient medical records were converted into HPO terms using Neji concept recognition^38^ with manual review by laboratory genetic counsellors or clinicians. Patients were selected for inclusion in this study based on having one or more HPO terms from a list of 716 that fell under “abnormality of the nervous system”^30^. The study was conducted in accordance with all guidelines set forth by the Western Institutional Review Board, Puyallup, WA (WIRB 20162523). Informed consent for genetic testing was obtained from all individuals undergoing testing, and WIRB waived authorization for use of deidentified aggregate data. Individuals or institutions who opted out of this type of data use were excluded.

The samples underwent exome sequencing as previously described^37^ with either SureSelect Human All Exon v4 (Agilent Technologies, Santa Clara, CA), Clinical Research Exome (Agilent Technologies, Santa Clara, CA), or xGen Exome Research Panel v1.0 (IDT, Coralville, IA). They were sequenced with either 2x100 or 2x150bp reads on HiSeq 2000, 2500, 4000, or NovaSeq 6000 (Illumina, San Diego, CA). Reads were mapped to the published human genome build UCSC hg19/GRCh37 reference sequence using the Burrows Wheeler Aligner (BWA) (using either v0.5.8 to v0.7.8 depending on the time of sequencing)^31, 32^. BAM files were then converted to CRAM format with Samtools version 1.3.1^39^ and indexed. Individual gVCF files were called with GATK v3.7-0 HaplotypeCaller^34–36^ in GVCF mode by restricting output regions to the RefGene primary coding regions +/- 50bp. Single-sample gVCF files were then combined into multi-sample gVCF files, with each combined file containing 200 samples. These multi-sample GVCF files were then jointly genotyped using GATK v3.7-0 GenotypeGVCFs. GATK v3.7-0 VariantRecalibrator (VQSR) was applied for both SNPs and indels, with known SNPs from 1000 Genomes phase 1 high confidence set and “gold standard” indels from Mills *et al.*^40^.

### Relatedness estimation

Relatedness between samples within each cohort was estimated using KING –kinship^41^. For DDD, we used common variants (MAF>0.01) that passed the following hard genotype filters: genotype quality (GQ) > 20, depth (DP) > 7, p-value from a binomial test on allelic depth > 0.001, and (after applying those genotype-specific filters) low missingness (<5%). For GeneDx, we used the same set of SNPs as used in the PCA, described in the next section. We used a cutoff of kinship coefficient > 0.04419417 to define related individuals, which is the lower bound cutoff for third-degree relatives. For each cohort, a list of unrelated parents and unrelated probands was created in a way that maximized the number of samples retained.

### Ancestry assignment

#### Assigning broad-scale genetically-inferred ancestry groups

To identify samples with similar genetic ancestry, we subset the genotypes of samples from 1000 Genomes phase 3^42^ and the Human Genome Diversity Project (HGDP)^43^ to the common SNVs (MAF>0.01) with low missingness (<10%). We ran pairwise LD pruning using plink^44^ (--indep-pairwise 50 5 0.2). We applied hard genotype filters to the DDD and GeneDx datasets (GQ>20, DP>7, and, subsequent to this, genotype missingness <10%), and took the intersection of the remaining variants across the datasets (N=17,693 SNVs). The first 20 PCs of the 1000 Genomes and HGDP reference cohorts were calculated using GCTA (version 1.93.0)^45, 46^. Using the SNP loadings, we project the DDD and GeneDx samples onto the reference sample PCs. Using the first seven PCs, we ran UMAP^47^ with the umap-learn python package with parameters - min-dist=0 and n_neighbours=100. This created the six clusters seen in **Supplementary Figure 3**, with the corresponding labels applied to DDD and GeneDx samples based on the locations of the reference individuals with known ancestry. We refer to these as “genetically-inferred ancestry (GIA) groups”.

#### Fine-scale ancestry

To assign fine-scale genetic ancestry to the individuals within each GIA group, we ran PCA on the individuals from 1000 Genomes and HGDP as well as the unrelated parents in the GeneDx dataset from that GIA group. We then projected the remaining samples from GeneDx and the DDD samples from that GIA group onto these PCs. To assign individuals to GIA sub-groups, we took the PCs that captured the majority of the variation within the broad-scale GIA group within which they fell, used these as input for UMAP, then ran HDBSCAN (a clustering algorithm)^48^ on the UMAP coordinates to create fine-scale clusters which we call GIA sub-groups (**Supplementary Figure 4**).

### Variant QC

Autosomal SNVs and indels underwent quality control (QC) separately within each cohort. In brief, we restricted to the variants within the intersection of the calling regions of the two cohorts, calculated metrics to determine the quality of SNV and indels passing a set of different thresholds, then selected the thresholds so as to optimise these quality metrics (**Supplementary Figures 5 and 6**).

The QC was conducted using bcftools version 1.16^39^.

#### SNV QC

The following genotype- and variant-level metrics were tested:

Genotype-level metrics:

1. Genotype Quality (GQ > {20,25,30})
2. Depth (DP > {7, 10})
3. Binomial p-value of allelic depth (for heterozygotes) (P(AD) > {0, 0.001})

Variant-level metrics:

1. VQSLOD (VQSLOD > {-1.5, -2.0, -2.5, -3.0})
2. Fraction of non-missing genotypes passing genotype-level QC thresholds (FPASS > {0.5, 0.7})

For each combination of metrics, we measured the following:

1. Transmission rate of synonymous singletons. Specifically, we identified synonymous variants seen in a single parent in the dataset, and determined what fraction of these were transmitted to the child. Since these variants are unlikely to contribute to the phenotype, we expect them to be transmitted 50% of the time, so we optimised the choice of QC parameters to get this metric as close as possible to 50% while simultaneously optimising the other metrics (**Supplementary Figure 5**).
2. Sensitivity to detect known *de novo* SNVs. Previous analysis had identified a total of 41,890 (38,038 SNV, and 3,852 indel) likely pathogenic *de novo* mutations across the two cohorts that passed stringent quality control^5^. We wanted to retain as many of these variants as possible (**Supplementary Figure 5**).
3. Number of variants that are called as homozygous for the alternate allele in the child and homozygous for the reference allele in both parents. These candidate *de novo* mutations are almost certainly errors.
4. Number of candidate *de novo* mutations that are seen in multiple individuals across the dataset (recurrent *de novos*), likely to be enriched for errors.
5. Rate of Mendelian errors in trios.
6. Total number of variants passing QC. We wish to maximise this while simultaneously optimising the other metrics. (**Supplementary Figure 5**)
7. The transition to transversion ratio Ts/Tv (**Supplementary Figure 5**).

We chose the following thresholds for SNV QC:

For DDD:

- GQ > 20
- DP > 7
- P(AD) > 0.001
- VQSLOD > -2.0
- FPASS > 0.5

For GeneDx:

- GQ > 25
- DP > 10
- P(AD) > 0.001
- VQSLOD > -2.0
- FPASS > 0.7

#### Indel QC

The following genotype- and variant-level metrics were tested:

Genotypes-level metrics:

1. Genotype Quality (GQ > {20,25,30})
2. Depth (DP > {7, 10})
3. Allelic Balance (Variant Allele Frequency) for heterozygotes (AB > {0.2, 0.3})

Variant-level metrics:

1. VQSLOD (VQSLOD > {-2.0, -5.0}, and no VQSLOD)
2. Fraction of genotypes passing genotype-level QC thresholds (FPASS > {0.5, 0.7}).

For each combination of metric we measured:

1. Transmission rate of rare inframe variants in low pLI, non-monoallelic DDG2P genes (**Supplementary Figure 6**). The logic here is the same as for the synonymous singletons mentioned above; inframe indels in these genes are likely under minimal selective pressure, on average, so we calibrated our QC so that the transmission of these variants was as close as possible to 50% while also optimising the other metrics. We also filtered to variants with MAF<0.001 in (or absent from) gnomAD^49^ for checking this metric.
2. Sensitivity to detect *de novo* indels. A total of 3,852 *de novo* indel mutations were detected across the two cohorts in previous analyses^5^. We wanted to maximise our sensitivity to detect these mutations (**Supplementary Figure 6**).
3. Number of coding indels passing QC (**Supplementary Figure 6**).
4. Ratio of frameshift to nonsense variants. From previous studies^50, 51^, we expect the ratio of frameshift to nonsense mutations to be roughly 1.2.

We chose the following thresholds for indel QC: For DDD:

- GQ > 20
- DP > 7
- AB > 0.2
- No VQSLOD
- FPASS > 0.5

For GeneDx:

- GQ > 30
- DP > 7
- AB > 0.3
- No VQSLOD
- FPASS > 0.7

The number of variants before and after QC is given in **Supplementary Table 3**.

### Sample QC

After applying the variant and genotype QC, we carried out sample QC by running regressions of different quality metrics on several covariates (detailed below), then removing individuals whose residuals were greater than four median absolute deviations from the median for these two regressions (**Supplementary Figure 8**).

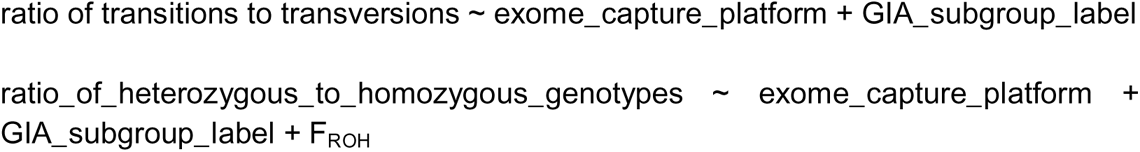

In addition, we removed individuals with a proportion of genotypes missing greater than 0.2. The number of individuals in each GIA sub-group before and after QC is given in **Supplementary Table 2**.

### Variant annotation and filtering

#### Initial variant filtering

We calculate the allele frequencies amongst unrelated, unaffected parents within all GIA sub-groups in our data that have 1150 unrelated, unaffected parents, from the two cohorts separately. We retained variants with MAF≤0.01 within all GIA sub-groups available, and with MAF<0.01 in all gnomAD v2.1.1 GIA groups^49^.

We removed any variants that overlap with a known recent segmental duplication^52^ or a simple tandem repeat^53^ obtained from the UCSC browser^54^. We also removed any variants which do not overlap the intersection of the bait regions from all exome captures used in the sequencing of both cohorts.

#### Variant annotation

Both cohorts were annotated using VEP v94.5^55^ including the LOFTEE plugin^49^. We focused on the annotation in the canonical transcript to group variants into three classes: synonymous, loss-of-function and functional.

As a control we considered synonymous variants with maximum SpliceAI^56^ score < 0.1.

Our classifications of loss-of-function (LoF) and functional variants were adapted from those used in gnomAD^49^ (https://github.com/broadinstitute/gnomad_methods/blob/main/gnomad/utils/vep.py). We classified the following VEP-predicted consequences as LoF, including only those that were predicted as high confidence (HC) LoFs by LOFTEE:

- splice_donor_variant
- splice_acceptor_variant
- stop_gained
- frameshift_variant
- stop_lost
- transcript_ablation

We grouped the following predicted consequences together into a group we call “functional variants”:

- missense_variant
- inframe_insertion
- inframe_deletion
- start_lost
- transcript_amplification
- protein_altering_variant
- splice_region_variant
- LoFs predicted to be low confidence (LC) by LOFTEE
- synonymous variant with a minimum SpliceAI score of 0.8

We removed variants if the predicted consequence in the canonical transcript did not fit into any of the categories listed above.

#### Filtering of missense and other functional variants

There are many metrics to predict deleteriousness of missense variants, but most of these are focused on predicting deleteriousness in the heterozygous state. We assessed several of these metrics (VARITY^57^, PrimateAI^58^, MPC^59^, CADD^60, 61^, ClinPred^62^, MoI-Pred recessive probability^63^, REVEL^64^, PolyPhen^65^), comparing the distributions for known pathogenic recessive missense variants in DDD (i.e. annotated as pathogenic/likely pathogenic in DECIPHER) to all missense variants on chromosome 20 (**Supplementary Figure 16**). We observed that PrimateAI and MPC were less discriminating than the other metrics in this context, and thus removed them. (Although CADD was also less discriminating, we retained it since it was available for all variants, unlike some of the other annotations.) We also removed VARITY-R since it uses the same model as VARITY-ER but just a different training set. For each of the remaining six metrics, we defined the threshold which gave us 90% sensitivity to detect the known pathogenic recessive missense variants (CADD_PHRED124.18; REVEL10.36; VARITYER_LOO10.25; PolyPhen10.59; ClinPred10.53; MoI-Pred recessive probability10.11). For inframe indels, we used a filter of CADD_PHRED117.34 (i.e. the CADD value which captures 90% of known inframe pathogenic recessive variants from DECIPHER^66^). For the remaining consequences that are included in the functional category, we use a filter of CADD_PHRED124.18.

We then evaluated the burden (observed/expected) and attributable fraction ([observed-expected]/sample size) (see section on Burden Analysis below) obtained for LoF/functional and functional/functional biallelic genotypes when requiring missense variants to pass the above cutoffs for different numbers of annotations (**Supplementary Figure 17**). Since not all of these deleteriousness metrics were available for all missense variants, we additionally evaluated the burden and attributable fraction when requiring missense variants to pass 170% of available annotations (**Supplementary Figure 17**). This final filter was the one chosen for the main analyses, on the basis of giving relatively high observed/expected (i.e. more significant enrichment) as well as relatively high attributable fraction (i.e. explaining more probands).

### ROH calling

Runs of homozygosity (ROHs) were called using bcftools-roh^67^ using common variants (MAF>0.01) with GQ120 and DP17 and low genotype missingness (<10%). Our previous work noted the effect of LD thinning on the calling of ROHs, with the optimal LD thinning differing by autozygosity levels^6^. We repeated the ROH calling for four values of LD thinning, r^2^={0.2,0.4,0.6,0.8}. The ROHs were called within each cohort for each GIA group independently to give a more accurate allele frequency estimate for the common variants used in the analysis. We retained ROHs that had quality score PHRED120. For each individual, we calculated the fraction of the genome in runs of homozygosity, F_ROH_. The distribution of F_ROH_ values for each GIA sub-group is shown in **Supplementary Figure 9**.

### Burden analysis

#### Sample filtering for the burden analysis

We removed trios in which both parents were inferred to come from different GIA sub-groups to the proband. Unless stated otherwise, we also removed trios with cross-continental admixture i.e. in which one parent was inferred to come from a different GIA group to the proband. **Supplementary Tables 4 and 6** also include results from sensitivity analyses in which we either a) removed all probands with any admixture (‘strict admixture filtering’ i.e. in which either parent was inferred to come from a different GIA sub-group from the child) or b) did no additional admixture filtering beyond requiring at least one parent to come from the same GIA sub-group as the child. We restricted all analyses to the twenty-two GIA sub-groups listed in **Table 1**, which were those that had at least 150 unrelated, unaffected parents, and a proportion of probands with cross-continental admixture <0.15. **Figure 1**, **Figure 2** and **Supplementary Figure 11** show exome-wide burden results for seven large GIA sub-groups combined (AFR4, AMR0, EUR4, EUR5, MDE3, SAS4 and SAS5), which were those that had at least 500 trios, for which the observed number of biallelic synonymous genotypes did not differ significantly from expectation (see below; unlike EUR1), and for which the exome-wide burden estimates were consistent when carrying out strict admixture filtering (unlike EUR7) (**Supplementary Table 4**).

#### Calculating the observed and expected number of biallelic genotypes

We extracted all observed trio genotypes with an observed rare allele in proband or parent to calculate the observed and expected number of biallelic genotypes. We removed genotypes within a trio if there was a Mendelian error or if any one of the three individuals had a missing genotype.

The expected number of biallelic genotypes per person was calculated in the same way as previously described in Martin et al.^6^ and summarised here. We considered four classes of biallelic genotype: LoF/LoF, LoF/functional, functional/functional and synonymous/synonymous. In short, the expected number of biallelic genotypes per person in GIA sub-group *p*, in variant class *c*, in gene *g* was calculated as:

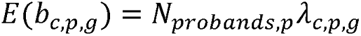

where *N_probands,p_* is the number of unrelated probands in GIA sub-group *p* and *λ_c,p,g_* is the expected frequency of biallelic genotypes given by,

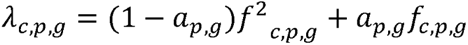

where *f_c,p,g_* is the cumulative frequency of parental haplotypes containing at least one variant of class *c* in gene *g* with MAF <0.01 in GIA sub-group *p*, and *a_p,g_* is the proportion of probands in GIA sub-group *p* with a ROH overlapping gene *g*. For the case of LoF/functional compound heterozygous genotypes, the expected frequency was calculated as:

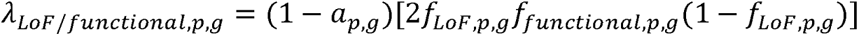

To calculate the cumulative frequency, we counted the number of haplotypes with at least one variant of class *c* in gene *g* in GIA sub-group *p* amongst unrelated, unaffected parents (*h_c,p,g_*), and divided this by the total number of haplotypes in that group (*N_haps_*).

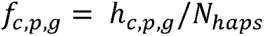

To calculate *h_c,p,g_*, within a gene, variant class, and GIA sub-group, for each unrelated, unaffected parent, we counted two haplotypes if a homozygous alternative genotype was observed, and one haplotype if a single heterozygous genotype was observed. When a parent had multiple heterozygous variants in that class within the gene, we tried to infer their phase based on transmission to the child, then counted one haplotype if they were in *cis* or two otherwise. If the phase was not clear, we counted one haplotype.

We determined compound heterozygous genotypes as those for which the proband inherited at least one heterozygous variant in the relevant class from each parent within a gene. To determine the observed count of biallelic genotypes, we counted the number of individuals with at least one homozygous alternative or compound heterozygous genotype within a variant class in the gene. If multiple deleterious biallelic genotypes were observed in a given individual, we only counted the one with the most severe consequence (e.g. if an individual had both a LoF/LoF and a LoF/functional compound heterozygous genotype in the same gene, this was counted only as LoF/LoF).

#### Testing for enrichment of biallelic genotypes over expectation

To determine the exome-wide burden of biallelic genotypes in variant class *c* in GIA sub-group *p*, we summed the observed and expected number across genes and compared these using a Poisson test. For the deleterious classes (LoF/LoF, LoF/functional and functional/functional) we used a one-sided Poisson test, testing whether the observed number was significantly greater than the expected, whereas for the biallelic synonymous class, we used a two-sided test. To determine the fraction of cases attributable to damaging biallelic genotypes, we calculated *O_c,p_* = ∑_g_ *O_c,p,g_* and *E_c,p_* = ∑*_g_ E_c,p,g_* for the three deleterious classes, then calculated the attributable fraction for GIA sub-group *p* as (∑_*c*_ *O_c,p_* - ∑_*c*_ *E_c,p_*)/*N_p_*, where *N_p_* is the number of unrelated probands in *p*. Within each GIA sub-group within each cohort, we chose the LD thinning threshold which gives us the maximum p-value in the synonymous variant class (given in **Supplementary Table 4**), then used this LD thinning threshold to calculate the *E_c,p,g_* for all variant classes for that GIA sub-group. We calculated the observed and expected values in DDD and GeneDx individually, and also conducted a pooled analysis by summing *O_c,p,g_* and *E_c,p,g_* across cohorts (**Supplementary Table 4**). The pooled estimates of attributable fraction calculated across the three deleterious genotype classes and across several GIA sub-groups (**Figure 1**, **Supplementary Figure 11**) are calculated as ∑_*p*_ [(∑_*c*_ *O_c,p_* - ∑_*c*_ *E_c,p_*)]/ ∑*_p_ N_p_* .

For our exome-wide burden analyses (**Figure 1**, **Supplementary Table 4**, **Supplementary Figures 10**, **11**, **12**, **15 and 17**), we removed genes flagged in gnomAD v2 as having an outlying number of synonymous variants, too many missense variants, or too many loss of function variants^49^, and genes that do not overlap with the intersection of the bait regions across all the exome capture kits used in DDD or GeneDx. This left 17,320 genes, of which 16,424 had at least one variant that passed our filtering.

In the Discussion, we present estimates of the residual risk of having an AR condition for undiagnosed patients without any candidate putatively damaging variants in known ARDD genes. To estimate this, we first removed individuals considered diagnosed and individuals with a damaging biallelic genotype that passed our filtering in a consensus or discordant gene. We then split the rest of the individuals into high (F_ROH_>0.0156) and low (F_ROH_<0.0156) autozygosity groups, and within each of these groups, calculated the attributable fraction in the genes not on the consensus or discordant gene lists.

#### Per-gene tests and multiple testing correction

For the per-gene enrichment tests, we initially tried implementing the original method used by Akawi et al.^8^, which is the exact probability for a sum of independent binomials. However, this method involves calculating all the possible ways the observed biallelic genotypes could have been distributed across the GIA sub-groups, and this proved to be computationally intractable for genes with high counts for our large sample size. Thus, we instead treated the total count of biallelic genotypes across GIA sub-groups as a sum of Poisson-distributed random variables with rates λ_1_, λ_2_,·, λ_n_ . This value follows a Poisson distribution with rate λ_1_ + λ_2_ +· + λ_n_ . Thus, we summed the observed and expected values across GIA sub-groups for a given gene and ran a one-sided Poisson test to determine the probability of observing at least ∑_p_ O_c,p,g_ genotypes given the expected number ∑_p_ E_c,p,g_. **Supplementary Figure 13** shows that this sum-of-Poissons approach gives very similar p-values to the previous approach (Pearson correlation R=0.98), particularly for genes with p<0.05 which are the ones of interest.

On each gene, we did four non-independent tests for these four likely damaging classes of variant:

- LoF/LoF
- LoF/LoF + LoF/functional
- functional/functional
- LoF/LoF + LoF/functional + functional/functional

As a Bonferroni correction we used p < 0.05/(17320 * 4) = 7.2 x10^-7^. We used estimates in all four damaging classes to calculate the Benjamini & Hochberg false discovery rate (FDR)-adjusted p-values. We also implemented a test based on synonymous/synonymous genotypes as a control, and reassuringly, the p-values from this followed the expected null distribution (**Supplementary Figure 13c**).

#### Burden explained by ClinVar pathogenic variants

Figure 2b shows the estimate of the AR attributable fraction after removing variants annotated as pathogenic/likely pathogenic (P/LP) in ClinVar. For this, we removed biallelic genotypes if the variant (for homozygotes) or both variants (in a compound heterozygote pair) fulfilled the following criteria:

- had CLNSIG=Pathogenic OR CLNSIG=Likely_pathogenic, OR
- had CLNSIG=Conflicting_interpretations_of_pathogenicity AND (CLNSIGCONF∼Pathogenic|Likely_pathogenic AND NOT CLNSIGCONF∼Benign|Likely_benign) i.e. if there were conflicting assertions of pathogenicity, at least one of those assertions was P/LP, but none were “benign” or “likely benign”

### Filtering and analysis of *de novo* mutations

*De novo* mutations (DNMs) were called by GATK Haplotype Caller, and variant calls were restricted to -/+ 50bp of RefGene primary coding regions. Specific filters for DNM calls were: calls were required to have greater than 10 reads in the proband, more than 3 supporting reads, a genotype quality of greater than 40 and a strand bias of less than 30, and allele fraction of >0.15 for SNVs and >0.25 for indels (except for calls on chrX in males, which were allowed to have an allele fraction of 1). Indels greater than 100bp and variants that were seen in more than 11 parents across the cohort were removed. Sites with an allele fraction of less than 0.3 were excluded if any one of the following conditions were met: BaseQRankSum (Z-score from Wilcoxon rank sum test of Alt Vs. Ref base qualities) <= 0.75, MQ (RMS Mapping Quality) <= 58 or QD (Variant Confidence/Quality by Depth) <= 8. Variants were annotated with bcftools-csq^68^ on the canonical transcript (Gencode GRCh38, version 43).

We calculated the expected number of DNMs in subgroupings of probands using a gene-specific null mutation rate model for different functional classes of mutations based on estimated triplet-specific mutation rates, accounting for gene length and sequence context^69^. The exome-wide attributable fraction was calculated as follows, for a given group of probands:

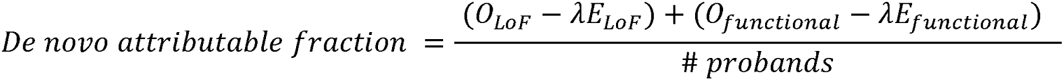

Where *O_c_* is the exome-wide observed number of *de novo* variants in consequence class *c*, *E_c_* is the expected number calculated using the model from^69^, and λ is a correction factor calculated as 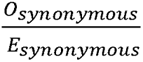.

#### Definition of known DD-associated genes

To get a list of “known” autosomal recessive developmental disorder-associated (ARDD) genes, we combined the list of genes from the Developmental Disorders Gene-to-Phenotype Database (DDG2P) used by DDD with a list of diagnostic genes used within the GeneDx in-house pipelines. There is an additional list of candidate genes in which GeneDx reports VUSs, but we do not consider these here. We downloaded the latest version of DDG2P on 6th March 2023 from https://www.ebi.ac.uk/gene2phenotype/downloads and retained those genes listed as having “definitive”, “strong” and “moderate” evidence (i.e. the clinically reportable categories) which were listed as “biallelic_autosomal”. This left 1,236 genes. From the GeneDx list (current July 2022), we retained those annotated as “validated” (i.e. the clinically reportable categories) which were listed as “autosomal recessive”. The GeneDx gene curation rules consider the following evidence in the course of validating a disease gene: 1) replication (at least two independent publications OR one large collaborative paper recruiting individuals from different backgrounds if GeneDx was involved) and 2) the number of probands segregating molecularly strong variants. There were 2,223 AR genes on the “validated” list, many of which are actually associated with disorders that are not DDs. Of these, we retained the 1,144 that were also “biallelic_autosomal” on the DDG2P list (at any confidence level), plus 331 which were not on the DDG2P ARDD gene list but which were classed as autism or intellectual disability genes by GeneDx. The remaining 748 genes were curated by two clinical geneticists (HVF and ES) to determine those that caused DDs as opposed to later-onset disorders, and of these, 191 were retained. The GeneDx ARDD gene list thus contained 1,666 genes. We called the 1,074 genes present on both the DDG2P and GeneDx ARDD gene lists “consensus” genes, and the 754 present on just one of those lists “discordant” genes; of these, 1,069 and 749 respectively were amongst the 17,320 genes retained for analysis (**Supplementary Table 5**).

### Phenotypic similarity of patients

The phenotypic similarity of patients was calculated following Kaplanis et al.^5^ with the *phenopy* package https://gitbub.com/GeneDx/phenopy. The pairwise similarity of two terms in The Human Phenotype Ontology (HPO) were compared quantitatively using the Hybrid Relative Semantic Similarity (HRSS) metric, and similarity for two lists of terms was calculated via a Best Match Average^21^. The phenotype similarity between two probands is defined as the listwise HRSS of the phenotypes describing each proband. Only terms currently in the HPO at the time of analysis were included, and any updates to retired HPO terms were handled by searching for the alternate IDs of all current phenotypes and replacing them where appropriate. The set of HPO terms assigned to each proband were pruned by removing ancestor terms in any ancestor-descendant pairs. As before, the information content (IC) of each term used for HRSS calculations was the mean of the IC based on the HPO-OMIM-ORPHANET phenotype-to-gene annotations and the phenotype-to-gene annotations of monogenic diagnosed cases from the relevant cohort/s (i.e. DDD alone if examining a pair of DDD patients, GeneDx alone if examining a pair of GeneDx patients, or DDD+GeneDx if examining a pair consisting of one patient from each cohort).

For the genes that passed FDR<5% in the genotype-based test, the phenotypic similarity of all pairs of probands with damaging biallelic variants in that gene were calculated. We compared the distribution of these scores to a null distribution of HRSS scores for 100,000 randomly-chosen pairs (**Supplementary Figure 14**). This null distribution was created such that the fraction of randomly-chosen pairs that involved a) two DDD patients, b) two GeneDx patients or c) one DDD and one GeneDx patient matched the fraction amongst the patients with damaging biallelic variants in the FDR<5% genes.

## Supplementary Note

### Phenotypic comparisons of the cohorts

DDD patients were slightly more male-biased than GeneDx patients (58.4% male versus 55.7% male, Fisher’s exact test p-value = 8x10^-8^). They were also slightly younger at recruitment on average (7.3 years versus 9.4 years; t-test p-value <1x10^-163^), and the age distribution was less variable (standard deviation 6.1 years for DDD versus 10.2 years for GeneDx) (**Supplementary Figure 1**). DDD patients had significantly fewer HPO terms on average than GeneDx patients (7.0 terms versus 19.8; t-test p-value<1x10^-200^). This likely reflects differences in how these HPO terms were recorded. For DDD, clinical geneticists recorded phenotypes that they thought likely to be relevant to a monogenic diagnosis and that were particularly distinctive amongst the population of rare disease patients being seen in genetics clinics, whereas in GeneDx, the HPO terms were extracted from the medical notes (including medical history and primary indication) through a mixture of automated text mining and manual curation by nurses, contractors and genetic counsellors trained in the abstraction process. Accordingly, there were multiple organ systems in which GeneDx patients were substantially more likely than DDD patients to have an HPO term, even after controlling for age and sex, including the musculature, digestive, cardiovascular, immune, respiratory and blood systems (**Supplementary Figure 2**). However, examination of the most common HPO terms revealed that many of those assigned to GeneDx patients are nonspecific (e.g. feeding difficulties, bruising susceptibility, failure to thrive) or represent common diseases (e.g. asthma, eczema) (**Supplementary Table 1**). These differences are likely related to differences in coding practices between the clinicians recruiting to DDD versus GeneDx rather than true phenotypic differences between cohorts.

### Suggestive new genes passing FDR<5% but not Bonferroni correction

Our signal in *ZDHHC16* was driven by one patient with a LoF/LoF genotype and two with LoF/functional genotypes (p=3.91x10^-6^). Two unrelated probands were strikingly similar phenotypically, having microcephaly, seizures, developmental regression/neurodegeneration, and abnormalities of the respiratory system. However, the third had a less distinctive phenotype involving generalised developmental delay with seizures, and had a sibling with a similar phenotype and the same *ZDHHC16* variants. Consistent with the neurodevelopmental features in these patients, *ZDHHC16* has been shown to play a critical role in the regulation of neural stem/progenitor cell proliferation in zebrafish telencephalic development^23^.

Biallelic variants in *HECTD4* (p=8.67x10^-6^ in our analysis) were recently reported to cause a neurodevelopmental disorder characterised by intellectual disability, seizures, movement disorder, behavioural abnormalities, macrocephaly, abnormality of dentition, and agenesis of the corpus callosum^24^. The two unrelated probands we observed with biallelic LoF/LoF genotypes in this gene also exhibited many of these features.

We found a single individual with a biallelic damaging genotype (homozygous LoF) in *ATXN1* (p=9.04x10^-5^), in which expansions of a trinucleotide repeat are a dominant cause of spinocerebellar ataxia type 1 (SCA1)^70^. There has also been a report of neurobehavioral abnormalities in the homozygous knockout mouse^25^. SCA1 is a neurodegenerative disorder rather than a developmental disorder, with typical onset in the third or fourth decade of life, but it can have early onset in childhood. The symptoms of our biallelic patient, who was recruited as a toddler, are consistent with SCA1, but there were additional clinical features including developmental delay and some dysmorphic features.

We observed three patients with damaging biallelic genotypes (all biallelic LoF) in *LRRC34* (p=5.42x10^-5^). A homozygous LoF in this gene has previously been reported in a patient with Joubert Syndrome, with accompanying functional evidence supporting pathogenicity ^26^. One of our three patients already has a monogenic diagnosis in a different gene, another had a phenotype consistent with Joubert Syndrome, but the third was phenotypically distinct. We consider that more evidence is required to definitively implicate *LRRC34* as a biallelic DD gene.

We observed two individuals with biallelic LoFs in *C11ORF94* (p=9.04x10^-5^), a known regulator of male fertility that plays a role in sperm-oocyte membrane binding^71, 72^. This gene has not previously been linked to developmental disorders. Our two patients did not have particularly similar phenotypes, and one had a partial diagnosis in another gene. Hence, we consider this finding very tentative and that more evidence is required to definitively implicate this gene as a DD-associated gene.

## Supplementary Figures

**Supplementary Figure 1:**
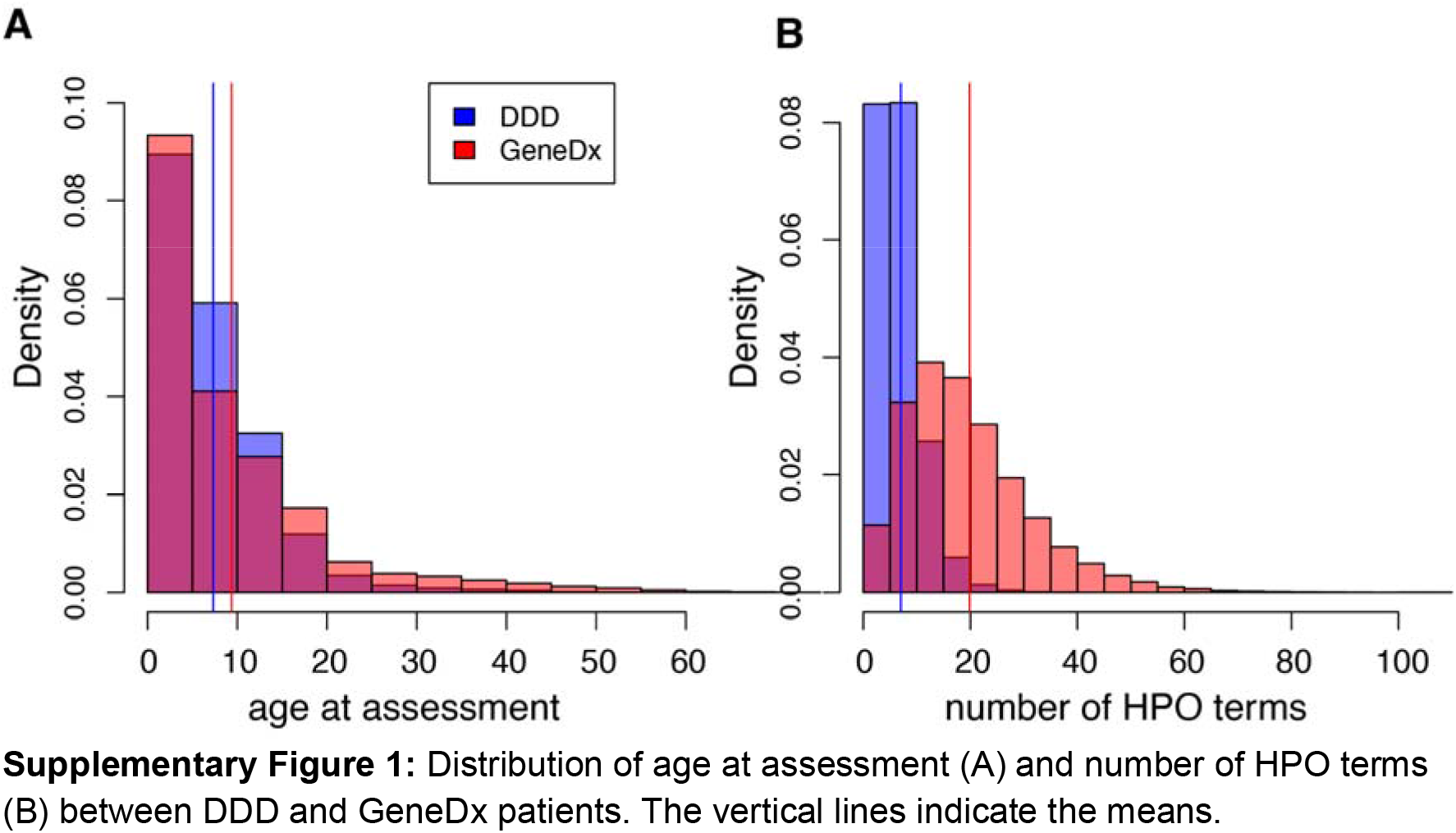
Distribution of age at assessment (A) and number of HPO terms (B) between DDD and GeneDx patients. The vertical lines indicate the means.

**Supplementary Figure 2:**
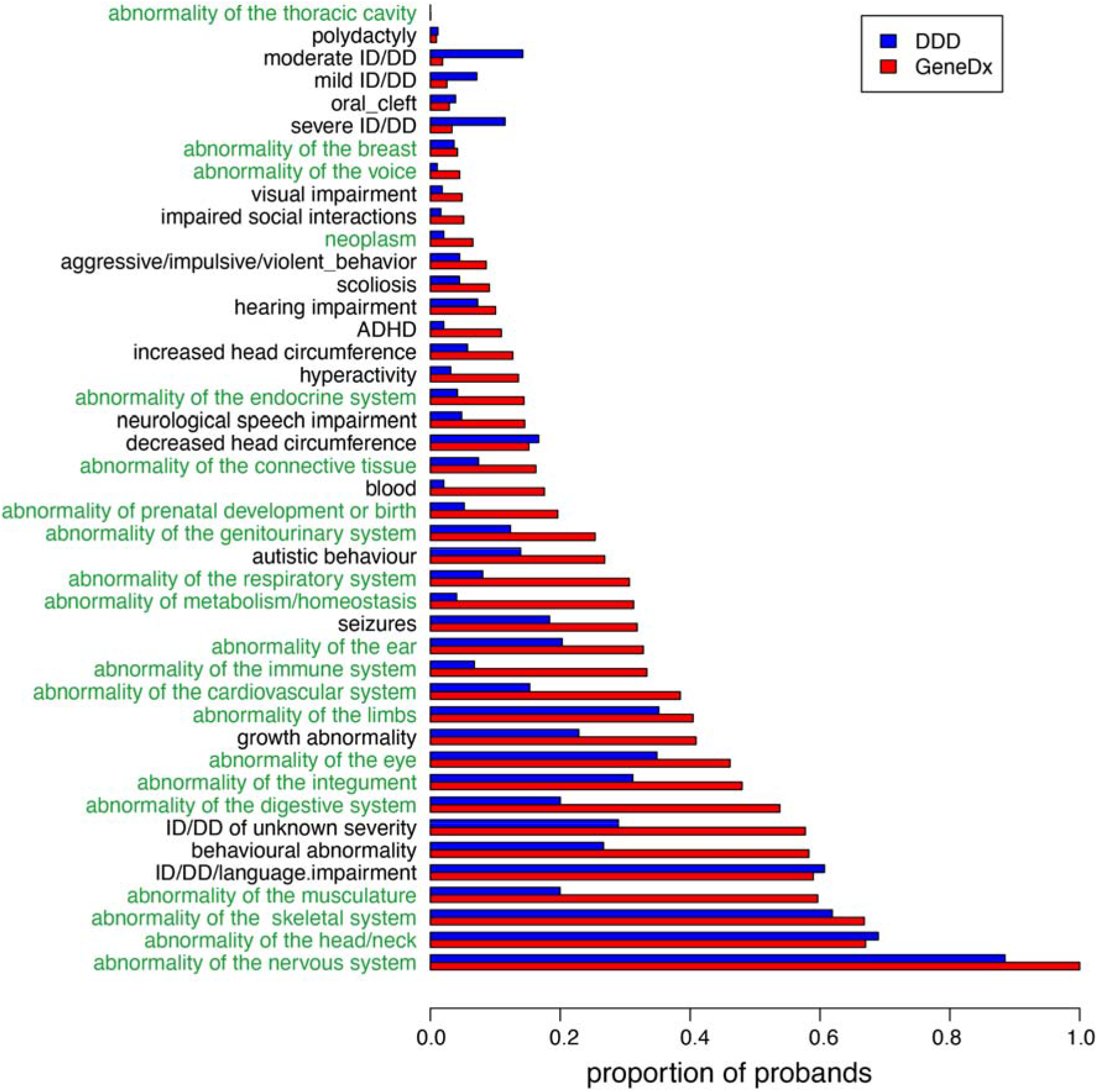
Proportion of probands from each cohort with at least one HPO term within the indicated chapter (green text) or specific phenotype (black text), ordered by the prevalence in GeneDx. We used logistic regression to test whether there was a significant difference in phenotype prevalence between cohorts after controlling for sex and age. All of the indicated phenotypes showed a significant association with cohort (p<0.0001) except the following: any ID/DD/language impairment, polydactyly, abnormality of the breast and abnormality of the thoracic cavity.

**Supplementary Figure 3:**
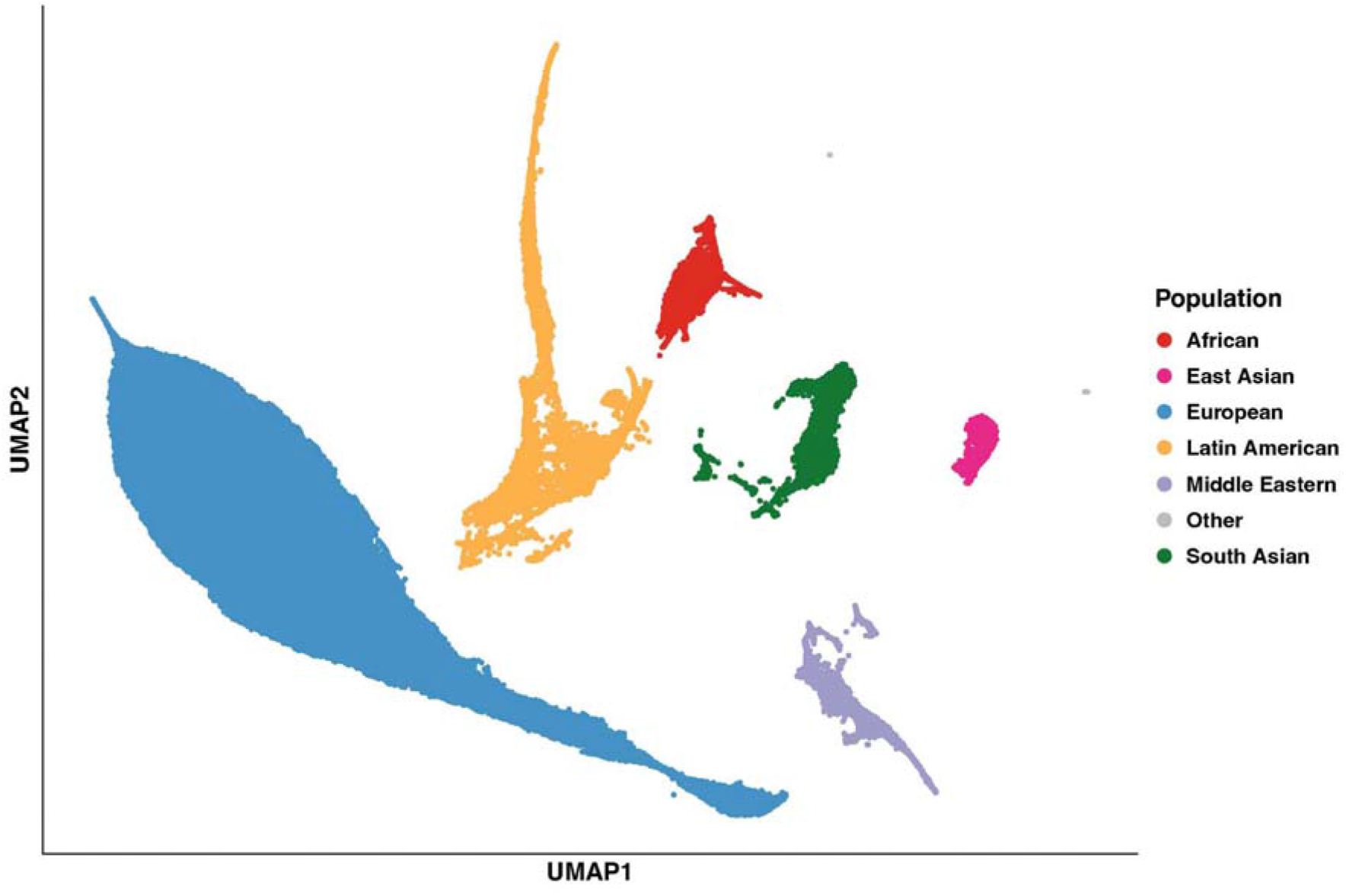
UMAP of the first seven principal components (PCs) of the 1000 Genomes and HGDP samples with DDD and GeneDx samples projected onto the PCs. The GIA groups were labelled based on the ancestry of the 1000 Genomes/HGDP reference samples within each cluster.

**Supplementary Figure 4:**
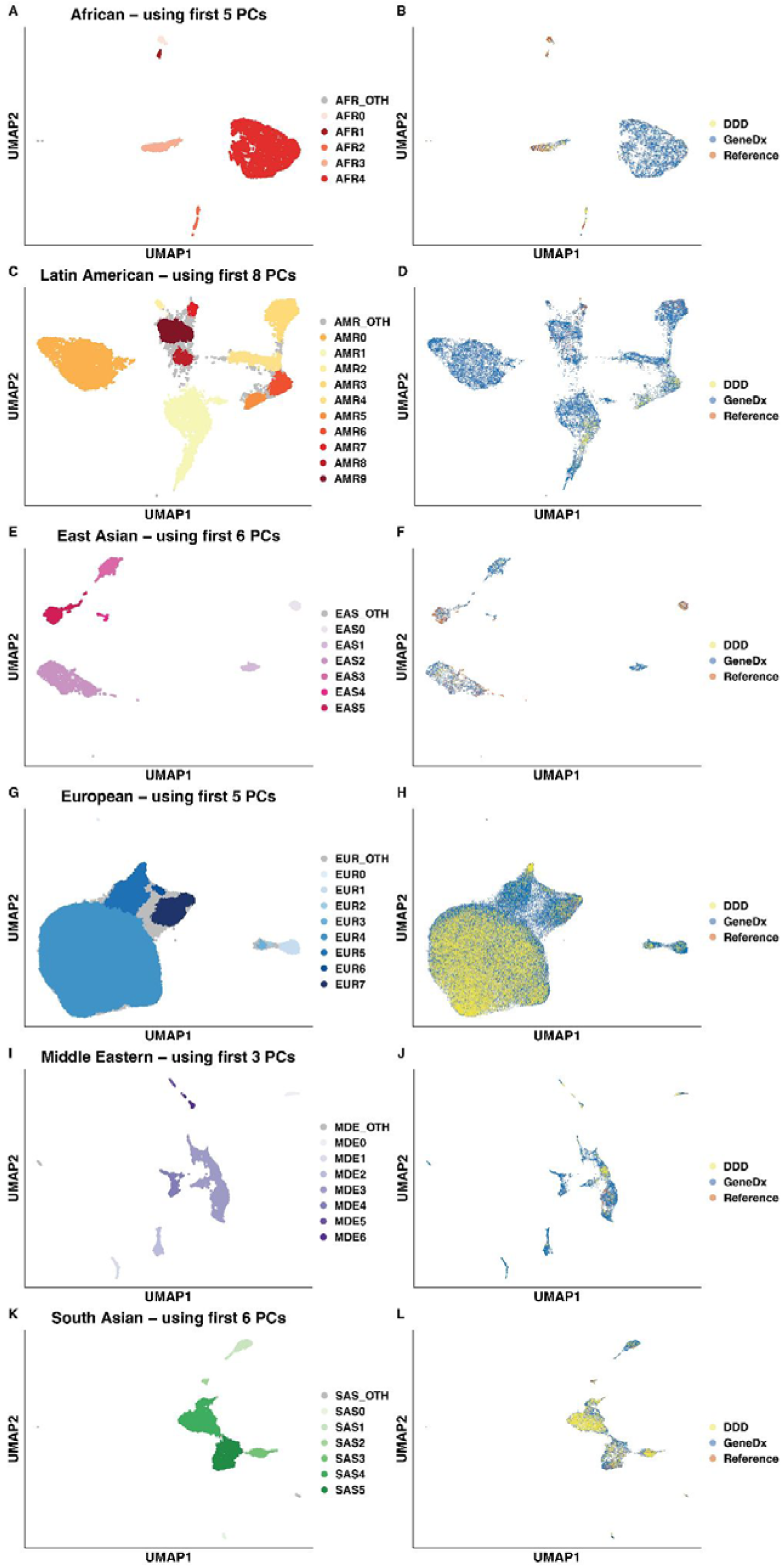
UMAPs based on principal components from each continental-level cluster. The PCA was run on each GIA group separately using the 1000 Genomes/HGDP reference samples together with the unrelated parents from GeneDx, then the DDD samples and remaining GeneDx samples were projected onto these. The clusters indicated in the left-hand plots were determined using HDBSCAN. The right-hand plots show the same UMAP but instead coloured to indicate which samples come from each cohort versus the reference samples. The GIA groups were as follows: A-B) African (AFR), C-D) Latin American (AMR), E-F) East Asian (EAS), G-H) European (EUR), I-J) Middle Eastern (MDE), and K-L) South Asian (SAS).

**Supplementary Figure 5:**
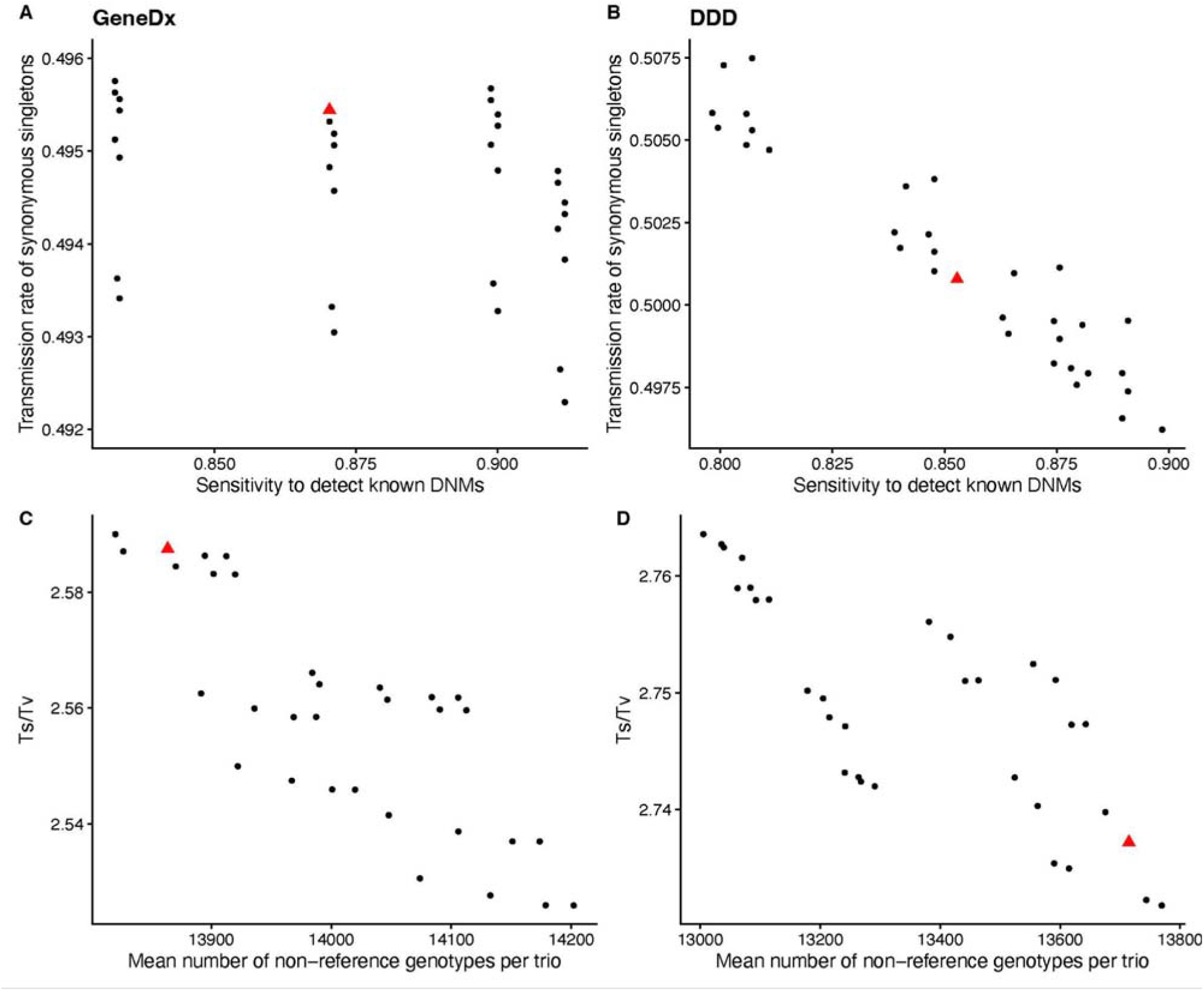
Choosing optimal QC metrics for SNVs. A) and B) show the transmission rate of synonymous singletons vs sensitivity to detect validated *de novo* variants in GeneDx and DDD respectively. Panels C) and D) show the transition to transversion ratio versus the mean number of non-reference genotypes per trio in GeneDx and DDD respectively. The red triangle represents the value for the final QC threshold chosen.

**Supplementary Figure 6:**
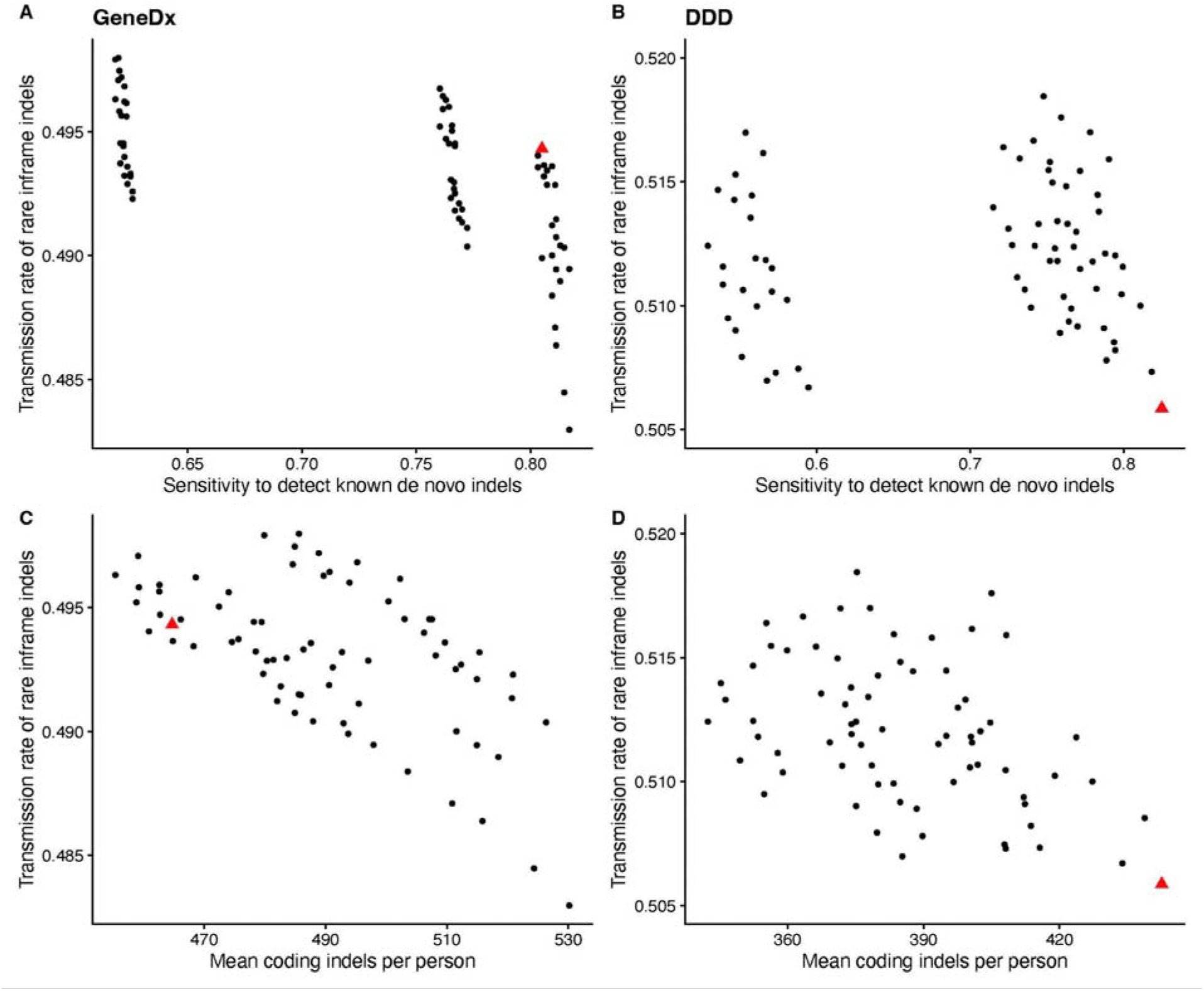
Choosing optimal QC metrics for indels. A) and B) show the transmission rate of rare inframe variants in low pLI non-monoallelic DDG2P genes vs sensitivity to detect validated *de novo* indels in GeneDx and DDD respectively. Panels C) and D) show the same transmission rate as the previous panels versus the mean number of coding indels per person in GeneDx and DDD respectively. The red triangle represents the final QC threshold chosen.

**Supplementary Figure 7:**
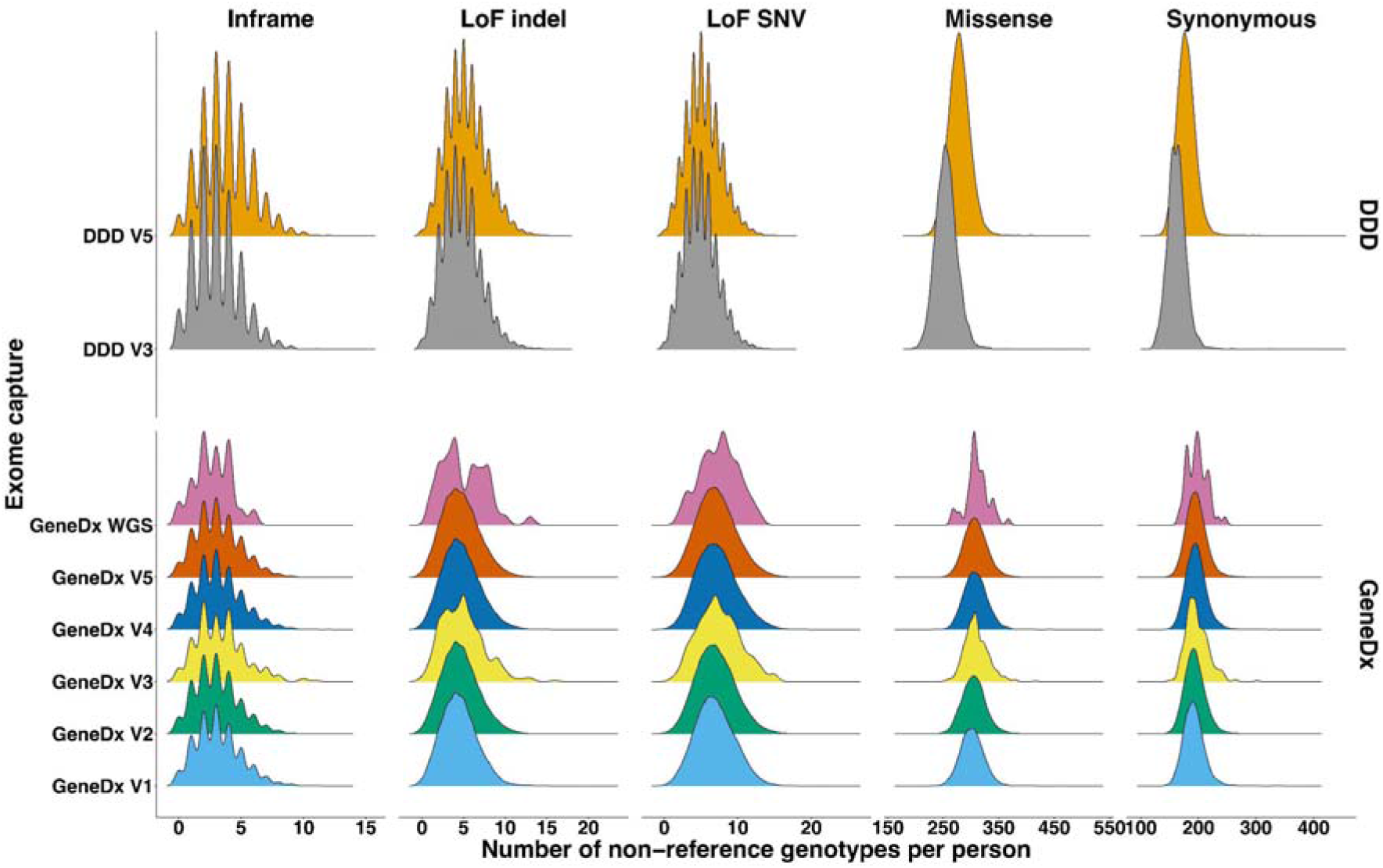
Rare variant (MAF<0.01 across the cohort) count distributions for different exome capture platforms for EUR4 individuals.

**Supplementary Figure 8:**
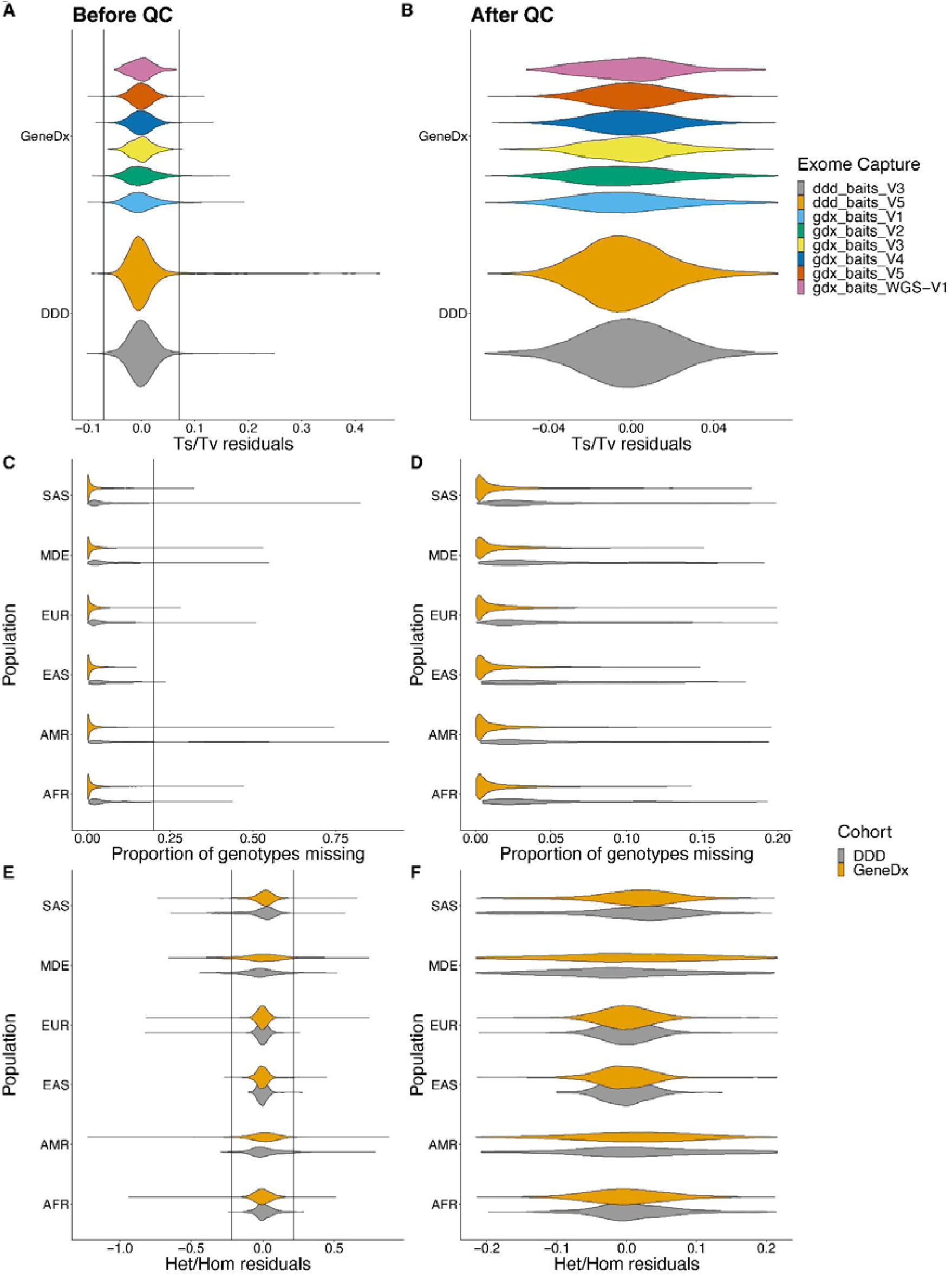
Distribution of various metrics per sample before and after sample QC. A) The residuals of the transition-transversion ratio, after regressing out the effect of population, and exome capture, split by exome capture before removing sample QC outliers and B) after removing the sample QC outliers. C) The proportion of genotypes missing, split by dataset, before removing sample QC outliers, and D) after removing sample QC outliers. E) The residuals of the heterozygous to homozygous alternative ratio, after regressing out the effect of population, exome capture, and F_ROH_, split by exome capture, before removing sample QC outliers, and F) after removing sample QC outliers. The vertical lines represent the thresholds for outlier removal (see Methods for details).

**Supplementary Figure 9:**
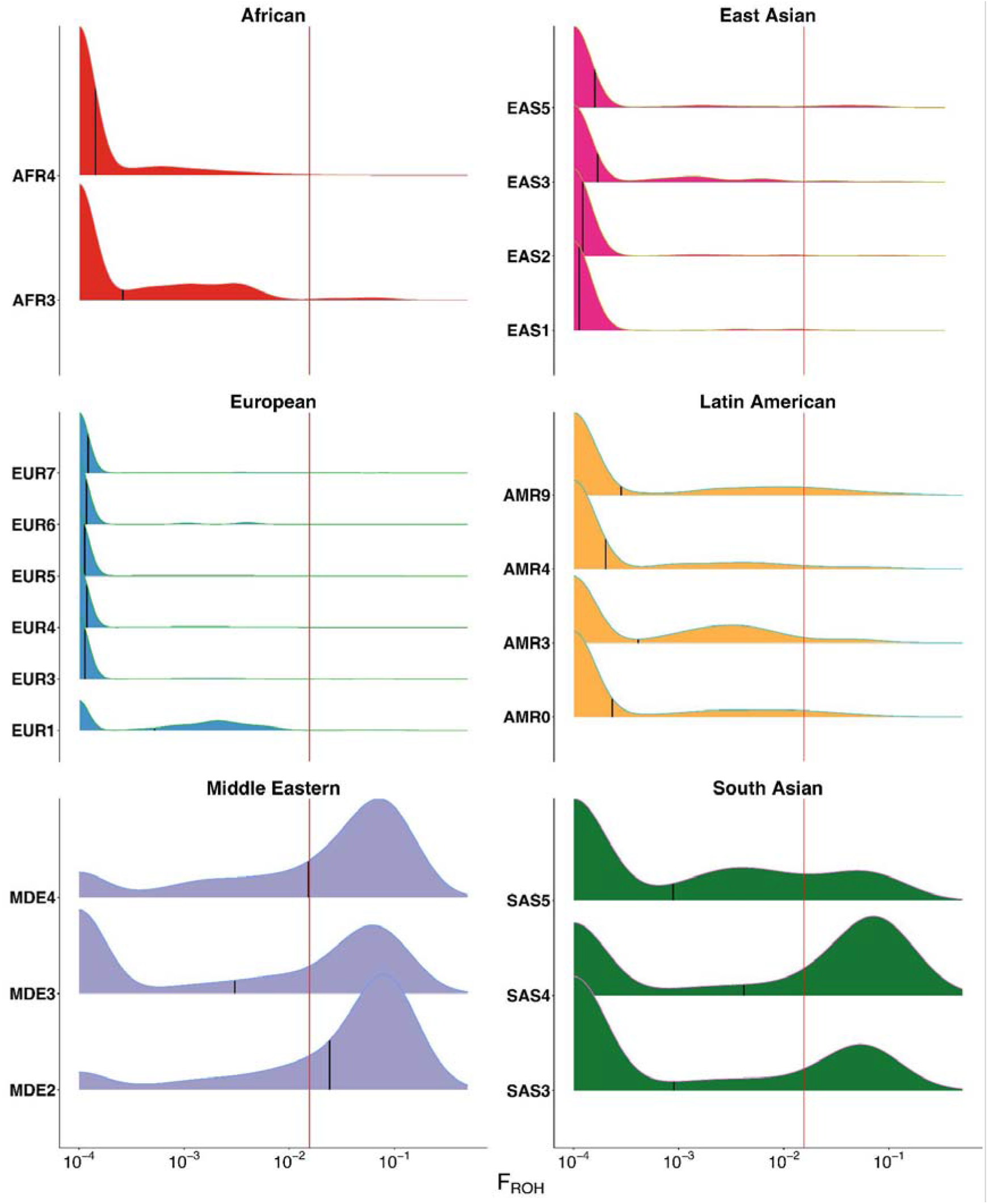
Density plots of distribution of F_ROH_ amongst the probands per GIA sub-groups on a pseudo-log scale (F_ROH_+0.0001). The vertical line represents F_ROH_=0.0156 which is the expected value for the offspring of second cousins.

**Supplementary Figure 10:**
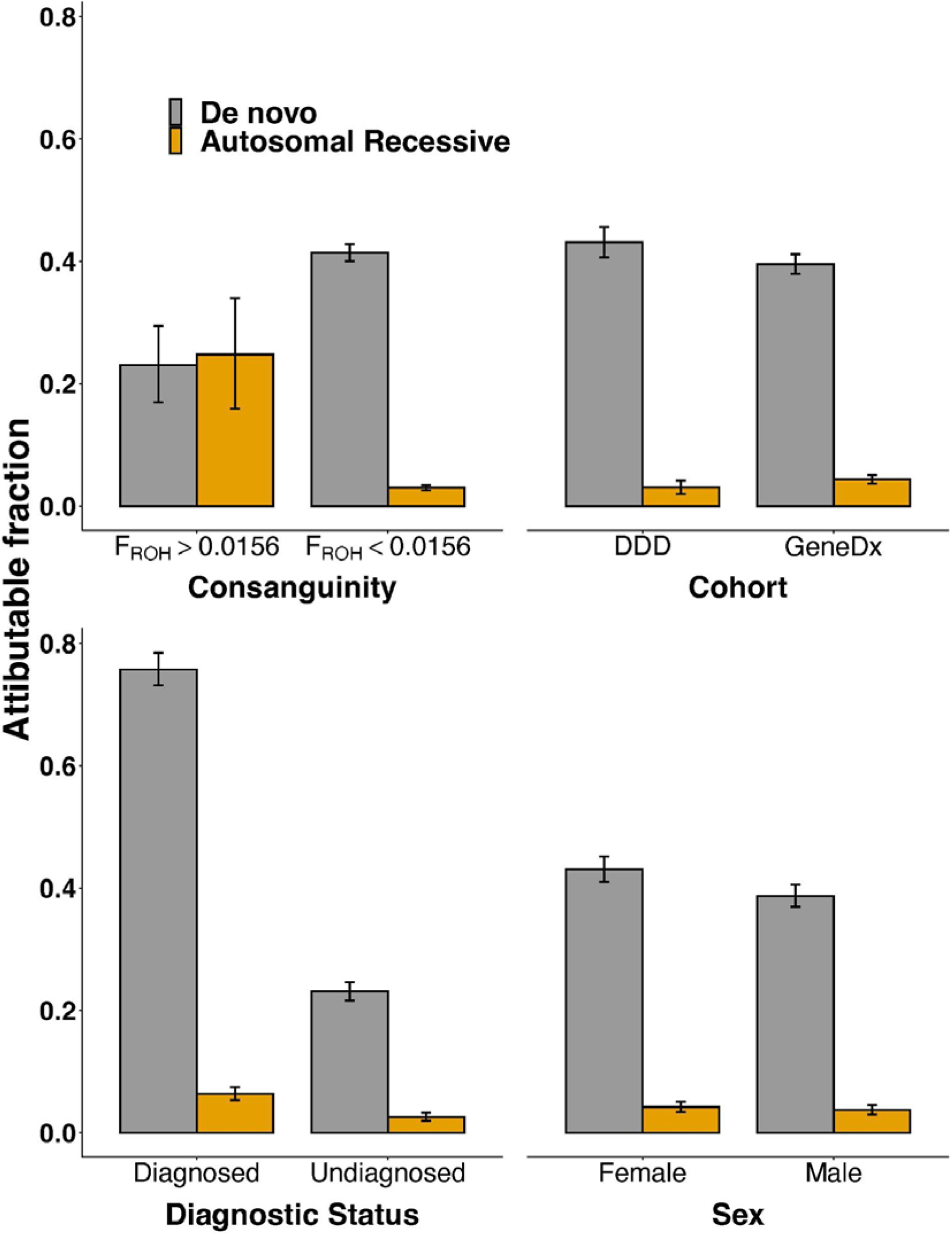
Exome-wide observed and expected number of biallelic genotypes per GIA sub-group, for the four consequence classes. This is after excluding trios with cross-continental admixture. This figure shows only GIA sub-groups with at least 200 trios; numbers for all GIA sub-groups are shown in Supplementary Table 4, together with estimates obtained with either no admixture filtering or stricter admixture filtering. The GIA sub-groups used in Figure 1 are shown in blue bold text along the x-axis. Coloured points are the observed numbers, black points are the expected numbers, and black lines show 95% confidence intervals around the observed. For some GIA sub-groups, the black points and/or black lines are not visible as they lie under the coloured points. P-values are shown for those where there is a nominally significant (p<0.05) difference between the observed and expected values, according to a Poisson test (two-sided for synonymous/synonymous, one-sided otherwise).

**Supplementary Figure 11:**
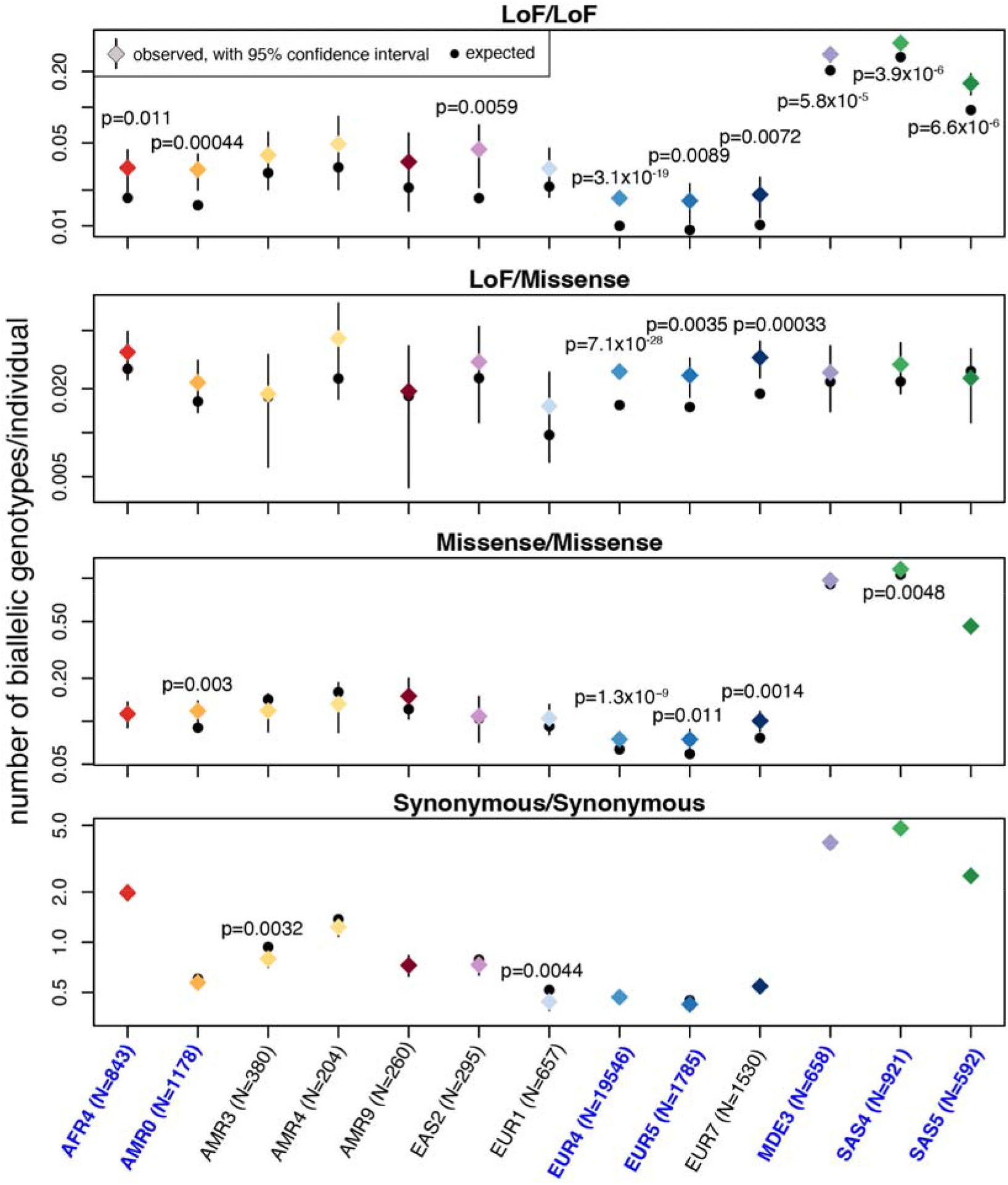
Fraction of patients in different groups attributable to *de novo* versus autosomal recessive coding variants [(observed-expected)/N]. The patients are split by level of consanguinity (A), cohort (B), diagnostic status (C) or sex (D). The bars show the overall estimates across the seven GIA sub-groups in Figure 1, excluding cross-continental admixed individuals.

**Supplementary Figure 12:**
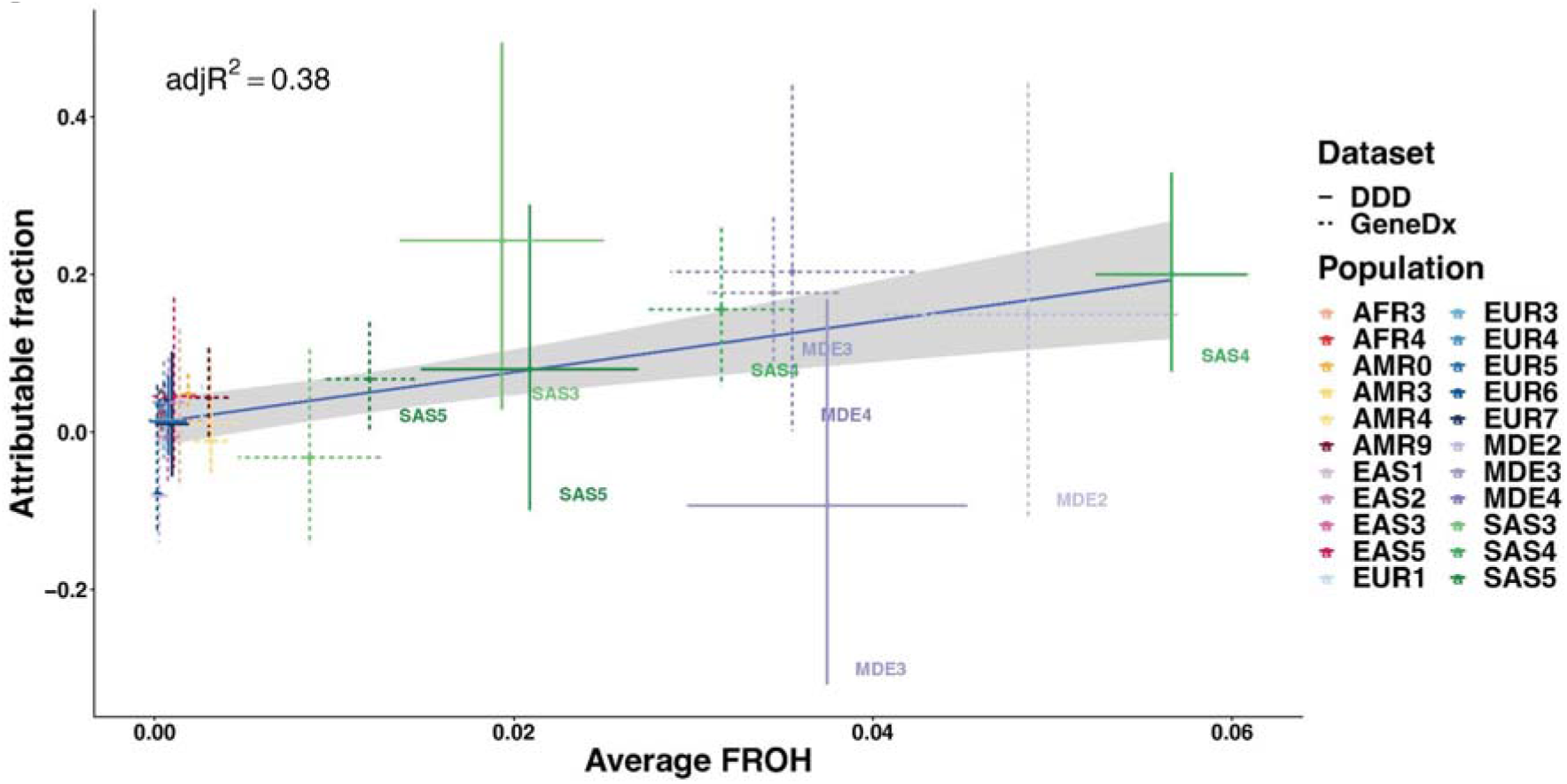
The estimated attributable fraction versus the average F_ROH_ for all twenty-two GIA sub-groups included in Table 1, split by cohort. The line of best fit is shown, with a 95% confidence interval around it shown in grey shading. The adjusted R^2^ is 0.38, significantly different from 0 (p=2.3x10^-4^).

**Supplementary Figure 13:**
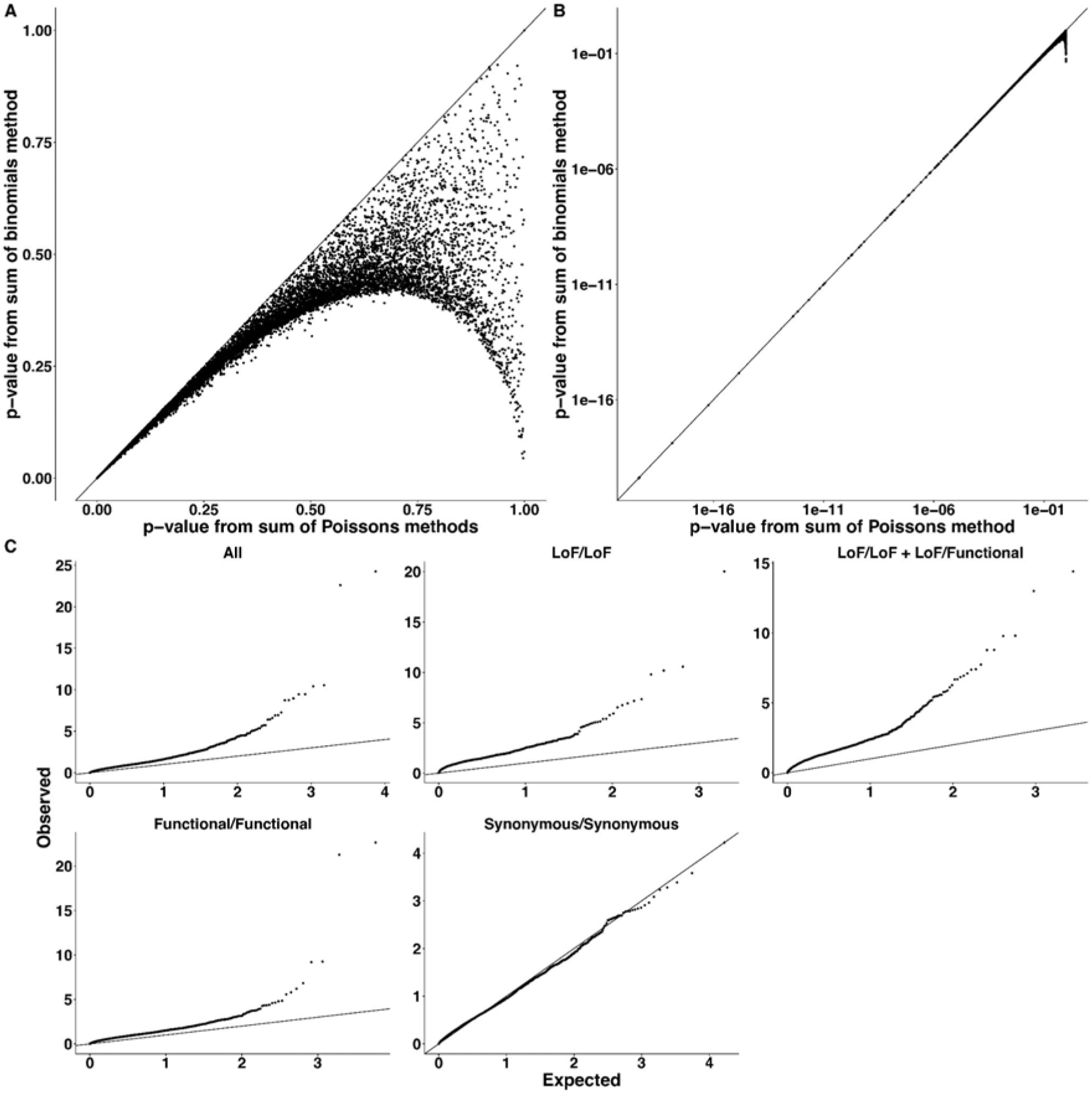
Performance of the per-gene tests. A) and B) are a comparison of the p-values, across every consequence class (i.e. four tests per gene plotted here), from the sum-of-Poissons method used in this paper with the sum-of-binomials method used in Akawi et al. ^8^ and Martin et al. ^6^. These show that the methods agree well for low p-values (Pearson correlation of 0.98 for panel A). Panel C) shows QQ-plots of the sum-of-Poissons method applied to 30,168 unrelated trios without cross-continental admixture from the twenty-two GIA sub-groups shown in **Table 1**, after removing genes with zero observed counts.

**Supplementary Figure 14:**
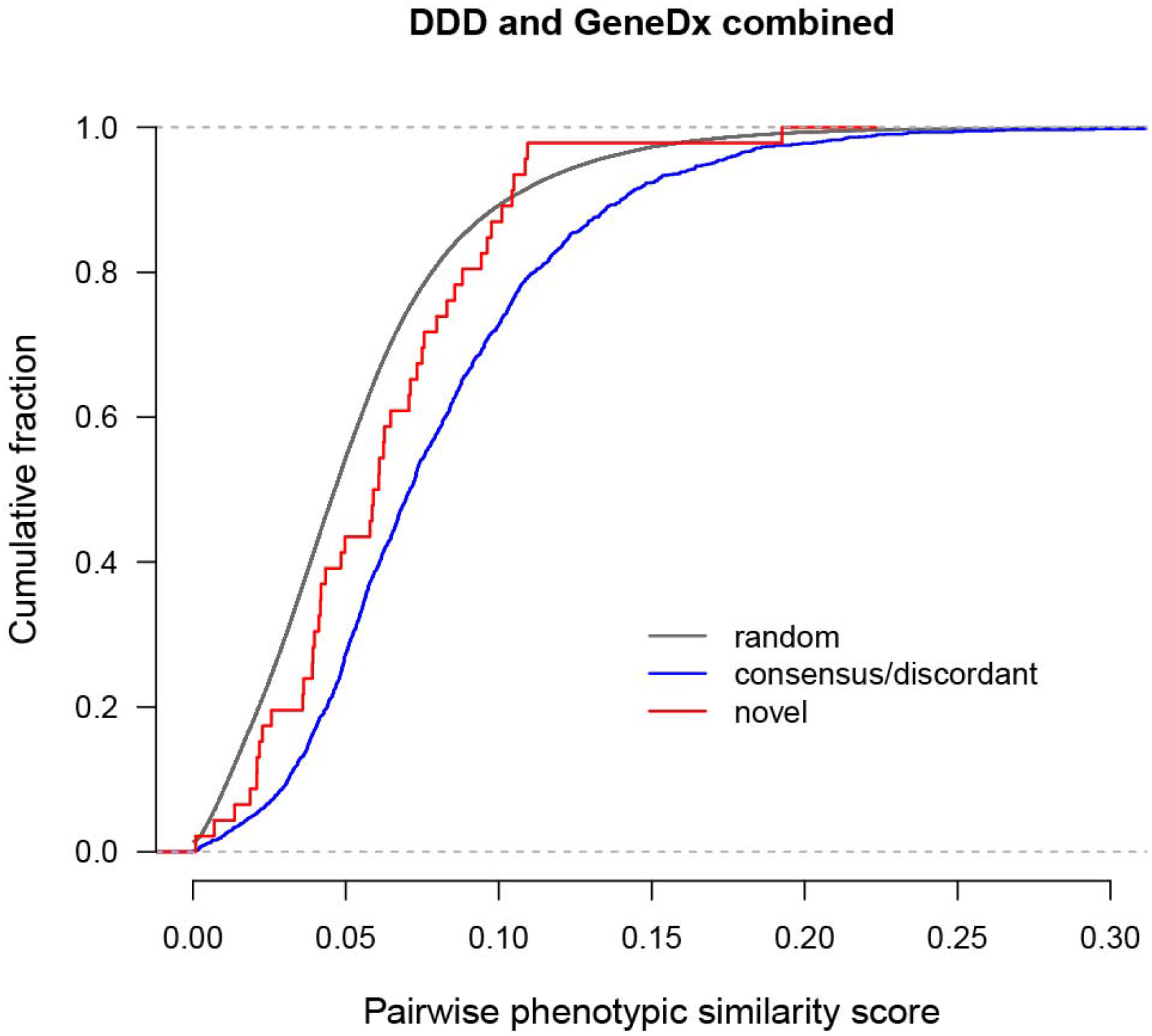
Cumulative distribution functions for pairwise phenotypic similarity scores as calculated by Phenopy. The distribution of novel genes (ATAD2B, ATG4C, ATXN1, C11ORF94, CRELD1, HECTD4, KBTBD2, LRRC34, ZDHHC16) passing FDR<5% is shown in red, consensus/discordant genes passing FDR<5% in blue, and the similarity scores of random pairs in grey. Random pairs were selected proportionally to match the occurrence of DDD/DDD, GeneDx/GeneDx and DDD/GeneDx pairs in the novel and consensus/discordant sets. The phenotypic similarity scores in patients with damaging biallelic genotypes in the novel genes were significantly lower than those for patients with such genotypes in consensus/discordant genes (one-sided Wilcoxon rank sum p=0.003), but they were significantly higher than random scores (one-sided Wilcoxon rank sum p=0.024)

**Supplementary Figure 15:**
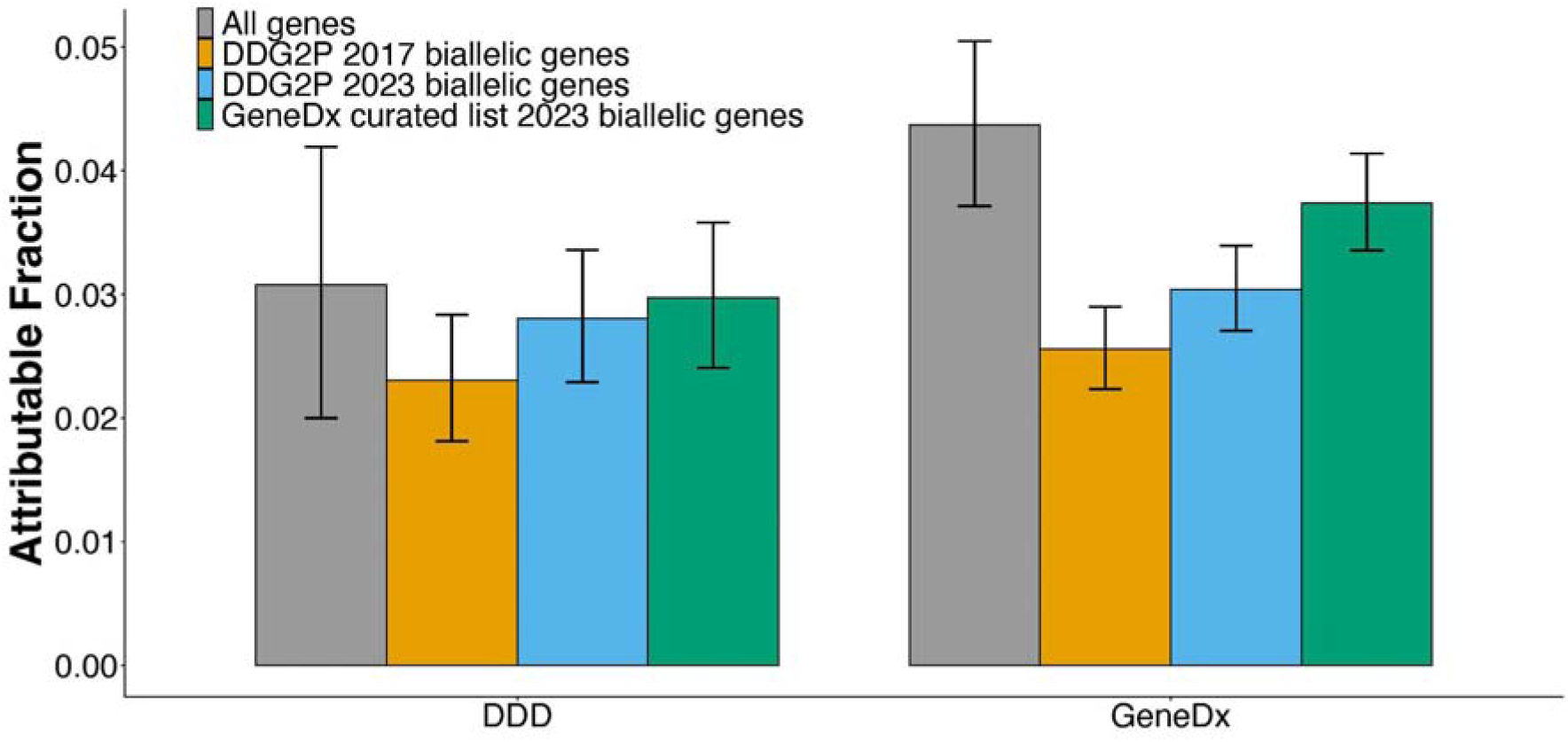
Fraction of patients in each cohort attributable to AR coding variants both across all genes and in the indicated ARDD gene lists.

**Supplementary Figure 16:**
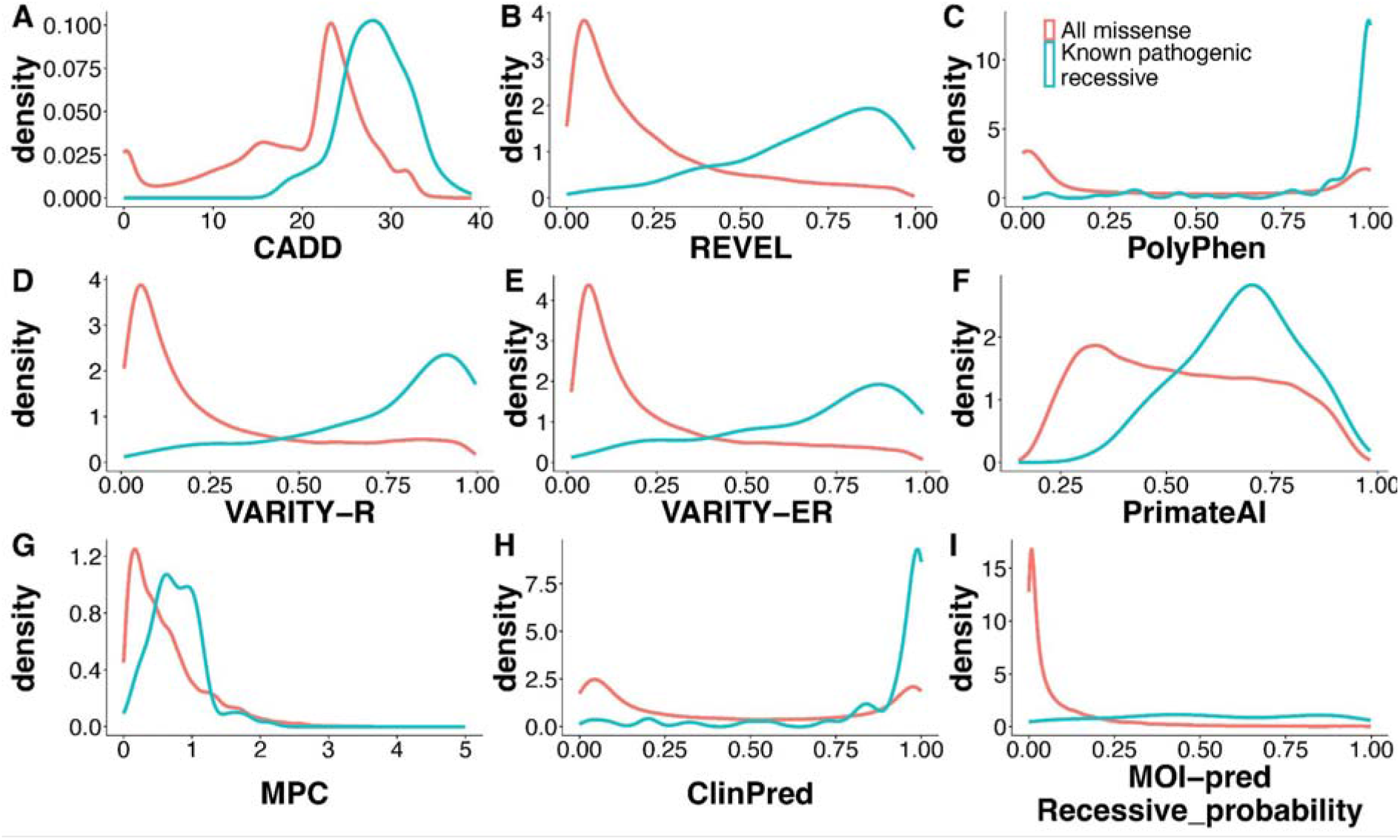
Distributions of pathogenicity predictors for 122 known pathogenic recessive missense variants from DECIPHER versus all missense variants on chromosome 20.

**Supplementary Figure 17:**
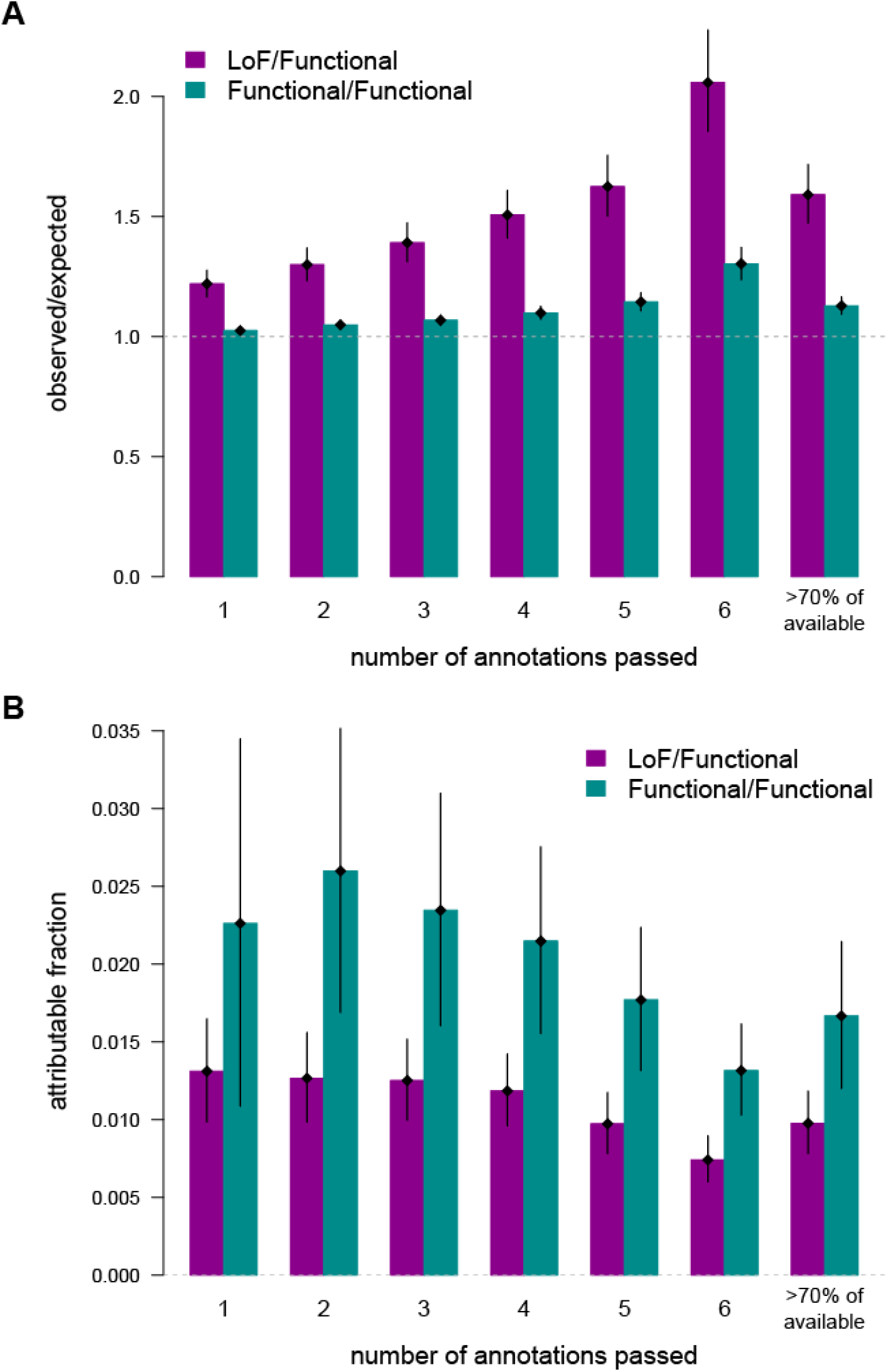
Effect of different strategies for filtering missense variants on exome-wide burden and attributable fraction. A) Ratio of observed to expected genotypes and B) attributable fraction [(Observed-Expected)/N], for LoF/functional and functional/functional genotypes, using different numbers of missense pathogenicity filters (see section on “Filtering of missense and other functional variants” in the Methods). Results are from the same samples as Figure 1 (i.e. the seven large GIA sub-groups with the cross-continental admixture filter, for GeneDx and DDD combined).

**Supplementary Figure 18:**
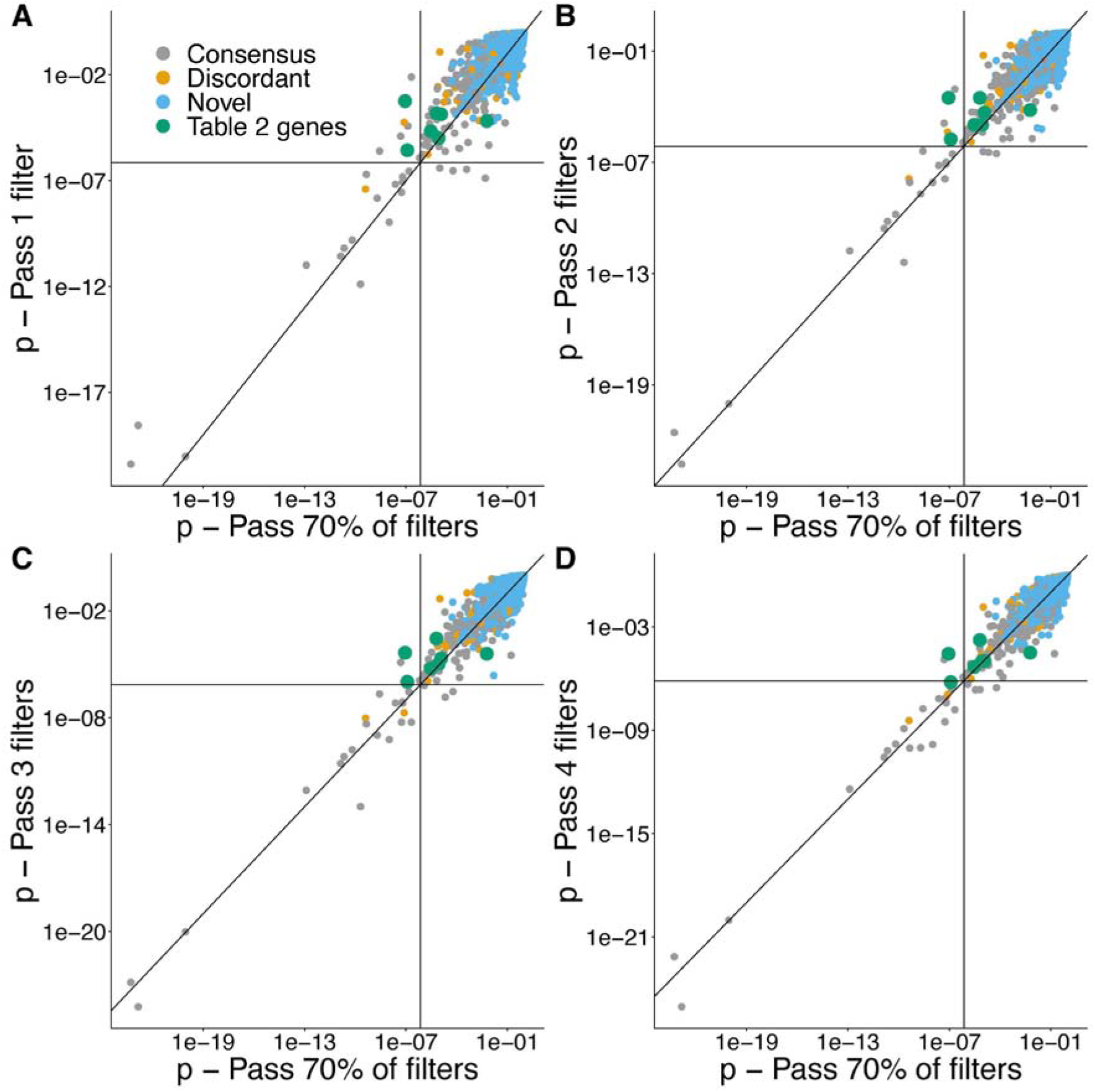
Comparison of p-values from the per-gene tests obtained using missense variants passing one, two, three or four deleteriousness filters versus passing 70% of missense deleteriousness filters (used in the main analysis). Only the p-value for the consequence combination that was most significant in the main analysis is shown. Genes highlighted in **Table 2** are coloured in green.

## Supplementary Tables

**Supplementary Table 1.** Comparison of prevalence of common HPO terms between 13,450 DDD patients and 36,057 GeneDx patients. Included are the HPO terms seen in >1% of patients from either cohort. The prevalence was compared using a Fisher’s exact test.

**Supplementary Table 2:** Sample sizes for each GIA sub-group, pre- and post-QC. This includes counts of all probands, probands in full trios (used in main analyses) with and without admixture filters, and of parents used for calculating the expected number of biallelic genotypes. The average F_ROH_ of full trio probands (post-QC) for each GIA sub-group is given, with and without admixture filters. The GIA groups labelled with a suffix “_OTH” are the individuals who were unassigned by the fine-scale ancestry classification; these probands were removed from all analyses unless stated. A total of 132 individuals that failed sample QC did not have any ancestry assignment in the GIA group assignment; these individuals are not included in this table or any genetic analyses.

**Supplementary Table 3.** Variant counts before and after QC for SNVs and indels for each cohort.

**Supplementary Table 4**. Results from exome-wide burden analysis. We show the results for DDD and GeneDx pooled, and each separately, with three different strengths of admixture filtering (1. cross-continental admixture removed, as used in Figure 1; 2. no admixture filtering; 3. all admixture removed), for the twenty-two analysed GIA sub-groups, as well as the totals across all GIA sub-groups and the seven large ones used in Figure 1. For each, we give the number of unrelated trios analysed (N), the observed (O) and expected (E) number of biallelic genotypes per consequence class, their ratio (O/E), the estimate of attributable fraction ([O-E]/N), a Poisson p-value for the difference between observed and expected (p_poisson), and the upper and lower bound of a 95% confidence interval around the observed value (lower and upper bound O 95CI). We also include the r^2^ threshold used for LD pruning prior to ROH calling in each GIA group.

**Supplementary Table 5:** List of consensus and discordant genes using DDG2P and GeneDx curated lists.

**Supplementary Table 6:** Results from the per-gene tests. We include only the counts for 16,424 genes with at least one variant observed that passed our filtering. We present results based on four different sets of trios:

a) Diagnosed+undiagnosed, removing those with cross-continental admixture (sample sizes for the twenty-two GIA sub-groups shown in Table 1)
b) Undiagnosed only, removing those with cross-continental admixture
c) Diagnosed+undiagnosed, no admixture filtering
d) Diagnosed+undiagnosed, removing those with any admixture

For all four sets of trios, we show the overall observed and expected number of biallelic genotypes, the Poisson p-value and FDR-adjusted p-value. For (a) we also include a breakdown of the observed and expected counts for each GIA sub-groups within each variant consequence class tested.

**Supplementary Table 7.** Variants and chapter-level HPO phenotypes for novel genes that were FDR<5% significant in our analysis. Patient GDX7 was the only patient with consent to provide specific HPO terms and details.

